# Rethinking the Human Resource Crisis in Africa’s Health Systems: Evidence across Ten Countries

**DOI:** 10.1101/2022.06.17.22276571

**Authors:** Ashley Sheffel, Kathryn G. Andrews, Ruben Conner, Laura Di Giorgio, David K. Evans, Roberta Gatti, Magnus Lindelow, Jigyasa Sharma, Jakob Svensson, Waly Wane, Anna Welander

**Author notes:** (corresponding author –).

## Abstract

Sub-Saharan Africa has fewer medical workers per capita than any region of the world, and that shortage has been highlighted consistently as a critical constraint to improving health outcomes in the region. This paper draws on newly available, systematic, comparable data from ten countries in the region to explore the dimensions of this shortage. We find wide variation in human resources performance metrics, both within and across countries. Many facilities are barely staffed, and effective staffing levels fall further when adjusted for absenteeism. However, caseloads—while also varying widely within and across countries—are also low in many settings, suggesting that even within countries, deployment rather than shortages, together with barriers to demand, may be the principal challenges. Beyond raw numbers, we observe significant proportions of health workers with very low levels of clinical knowledge on standard maternal and child health conditions. This work demonstrates that countries may need to invest broadly in health workforce deployment, improvements in capacity and performance of the health workforce, and on addressing demand constraints, rather than focusing narrowly on increases in staffing numbers.

**Key messages:**

- This study analyzed health worker surveys from ten countries in Sub-Saharan Africa for a deeper understanding of human resource challenges.
- Average staffing across facilities is far below the stated staffing norms for each country.
- Half of health centers and health posts have one or fewer clinical staff assigned to them.
- Staffing is even lower when adjusted for absence, which is highest in small facilities and public facilities.
- Massive within-country variation in caseload suggests that staffing problems may be solved in part by reallocation of clinical staff.
- Health workers lack basic clinical competencies, caseloads are imbalanced, and there is substantial absence of workers from health facilities.

## 1. Introduction

For decades, experts both on the African continent and those in the international community have highlighted a shortage in human resources for health in Africa (Huddart and Picazo, 2003, World Health Organization, 2006, Gomes Sambo, 2005, Elkhalifa, 2014, Fieno et al., 2016). One analysis projected a shortfall of six million health workers across the continent by 2030 (Tulenko, 2016). African countries on average have fewer medical workers per capita than any other region in the world.^1^ Yet while a shortage of health workers is undoubtedly a concern, it is not the only human resource challenge to Africa’s health systems. Recent analysis has highlighted others, including low quality of health worker knowledge (Di Giorgio et al., 2020, Okeke, 2021) and low access to and utilization of health services (Abekah-Nkrumah, 2019). Complicating the debate is the fact that all of these factors are endogenous: understaffed or under-skilled health facilities may result in low service utilization, and low service utilization may affect the number of staff assigned to a facility.

Much of the literature on the crisis in human resources for health relies on either broad counts of health workers at national levels combined with population numbers or richer analyses that are focused on a single country. Previous analyses have focused on identifying cross-country gaps in health workforce throughout the world (WHO, 2006). Others have focused on the health workforce challenges unique to individual country contexts. For example, Okeke (2021) experimentally tests whether staffing shortages or staff ability is the binding constraint in Nigeria, finding evidence in favor of ability; and Fieno et al. (2016) examine the political economy of staffing challenges in Ethiopia, showing that with political will, strong state capacity, and additional resources it is possible to increase the supply of health workers. However, few analyses look more comprehensively at staffing shortages in Africa and in low- and middle-income countries by providing comparable analysis across countries that highlights both high-level country averages and within-country variation.

This paper draws on newly available data from ten countries in Sub-Saharan Africa to answer four key questions. First, how many health workers are there? Second, are health workers present and what are the implications of worker attendance on effective staffing levels? Third, how heavy are health workers’ caseloads? Fourth, how competent are health workers? We use the Service Delivery Indicators (SDI) surveys—facility surveys that are designed to be nationally representative and to provide comparable data across several countries—to demonstrate how challenges tend to be either consistent or divergent across settings. In this study, we focus on within-country description and cross-country comparison to answer these staffing questions and consider the implications for effective health care delivery in low- and middle-income settings across Africa.

## 2. Methods

### 2.1. Data: The Service Delivery Indicators

The SDI health surveys, led by the World Bank, are health facility assessments used in low- and middle-income countries (LMICs) to generate nationally representative data on health service delivery with a focus on primary health care service quality (Bold et al., 2011, Bold et al., 2010). Using standardized instruments adapted to each country context, the SDI collects both facility and health worker level information that includes service availability and readiness, health worker capacity to diagnose and treat common illnesses, and health worker presence through unannounced visits. Data are made publicly available by the World Bank through the SDI data repository (The World Bank, 2021).

This study draws on data from the Sub-Saharan Africa region: Kenya (2018), Madagascar (2016), Malawi (2019), Mozambique (2014), Niger (2015), Nigeria (2013), Sierra Leone (2018), Tanzania (2016), Togo (2013), and Uganda (2013). The SDI country reports contain comprehensive information on each survey’s methodology and questionnaires. Briefly described, all surveyed facilities completed a facility inventory questionnaire. In addition, a health worker roster collected information on the total number of health workers at the facility and was used to randomly sample up to ten health workers for follow-up on a second unannounced visit to the facility to assess health worker absenteeism. Clinical vignettes were presented to up to ten randomly selected health workers who conducted outpatient consultations at the facility.

The total study sample includes interviews from 79,441 health workers^2^, observations of absenteeism from 35,896 health workers, assessments of competency from 15,207 health workers, and audits from 8,916 health facilities from across ten countries in Africa. Details of the sample are listed in *Table 1* with further disaggregation by country, facility type, managing authority, urban/rural, and health worker cadre in *Supplementary Table S1*.

**Table 1:** Total number of facilities, health workers, health workers assessed for absenteeism, and health workers assessed for competency, by country.

| Country | Year | Total number of facilities | Total number of health workers - staffing | Total number of health workers - absenteeism | Total number of health workers - competency |
| --- | --- | --- | --- | --- | --- |
| Kenya | 2018 | 3,038 | 24,404 | 12,266 | 4,485 |
| Madagascar | 2016 | 444 | 2,200 | 1,565 | 619 |
| Malawi | 2019 | 1,106 | 13,290 | 4,305 | 1,522 |
| Mozambique | 2014 | 195 | 2,972 | 936 | 694 |
| Niger | 2015 | 255 | 1,331 | 661 | 514 |
| Nigeria | 2013 | 2,385 | 21,318 | 9,764 | 5,014 |
| Sierra Leone | 2018 | 536 | 5,055 | 2,087 | 829 |
| Tanzania | 2016 | 383 | 5,160 | 2,119 | 498 |
| Togo | 2013 | 180 | 1,364 | 917 | 303 |
| Uganda | 2013 | 394 | 2,347 | 1,276 | 729 |
| **Total** |  | **8,916** | **79,441** | **35,896** | **15,207** |

### 2.2. Empirical strategy

We estimated three dimensions of health workforce performance—availability, productivity, and competency—each necessary to the achievement of improved service delivery and health outcomes (World Health Organization, 2006). To assess the availability of health workers we examined staffing levels and health worker absenteeism. Specifically, to assess staffing, a complete listing of staff working at the health facility, including each individual’s cadre was collected through a staff roster questionnaire. Absenteeism was defined as the proportion of randomly selected health workers who were absent from the health facility during an unannounced facility visit who were scheduled to be present at the facility. We calculated total clinical staff adjusted for absenteeism by taking the total number of clinical staff from the roster and reducing that number by the proportion of clinical staff absent, based on the results of the absenteeism assessment.^3^

To assess heath worker productivity, we defined caseload as the number of outpatient visits recorded in outpatient records in the three months prior to the survey, divided by the number of days the facility was open during the three-month period and the number of health professionals who conduct outpatient consultations. In some analyses, this indicator was adjusted for the average absenteeism at the facility-level.^4^

To assess health worker competency, we examined diagnostic and treatment accuracy using data collected through clinical vignettes which are standardized clinical case simulations developed to assess the health workers’ ability to diagnose and treat common outpatient conditions (Das et al., 2008). Diagnostic accuracy was calculated as the percentage of vignettes for which the health worker gave the correct diagnosis while treatment accuracy was calculated as the percentage of vignettes for which the health worker gave the correct treatment. A combined measure of diagnostic and treatment accuracy was calculated as the percentage of vignettes for which the health worker gave the correct diagnosis and the correct treatment. Supplementary Table S2 provides a detailed description of the vignette requirements by condition used to calculate correct diagnosis and correct treatment.

Several additional facility level and health worker level measures were included in the exploration of the extent to which accounting for health worker characteristics explains differences in health worker performance. Infrastructure availability was calculated as the availability of three components: improved water source, improved sanitation, and electricity. Equipment availability was calculated based on four items being available and functional: a thermometer, a stethoscope, a sphygmomanometer, and a weighing scale. Detailed definitions for each measure can be found in Supplementary Table S3. Mean absenteeism and mean provider competency were calculated at the facility level for use in the facility level regression model. Facility type was reviewed across countries and reclassified into three categories (hospital,^5^ health center,^6^ and health post)^7^ while managing authority was reclassified into two categories (public and private/NGO).Similarly, health worker cadre was recategorized into three categories (doctors/clinical officers, nurses/midwives, and other)^8^ and health worker education into three broad categories (primary, secondary, and post-secondary education).

Descriptive results, summarized as proportions and means, were calculated for all main measures, and disaggregated by facility type, managing authority, and urbanicity where applicable. For health worker level analyses, results were also disaggregated by health worker cadre. For all analyses, survey weights were incorporated to account for the complex survey design.^9^ Descriptive analyses are presented separately for each country. In addition, we present the cross-country average calculated as the average of the ten country estimates. We investigated the association of our key health workforce measures (health worker absenteeism, caseload, and diagnostic and treatment accuracy) with facility and health worker characteristics using multivariable linear and logistic regression models with standard errors adjusted for clustering of health workers within facilities. The regression analysis was conducted on a pooled dataset of all countries except Uganda which was omitted due to missing data on key regression variables^10^. All analyses were conducted with Stata 15 and R-4.0.3 (StataCorp, 2017, R Core Team, 2022).

### 2.3. Ethical approval

Each of the SDI surveys were carried out in collaboration with the Ministry of Health in each country. All survey participants consented to participate in the survey. Details regarding the ethics approval are available in each country survey report. This is a secondary analysis of SDI data.

## 3. Results

### 3.1. How many health workers are there?

To understand how many health workers there are, we first looked at the distribution of health worker cadres by facility type (Table 2). We then looked at staffing patterns to understand the average number of clinical staff per facility as well as the proportion of facilities with limited clinical staff present (Table 3). We compared those levels with staffing norms for the countries (Supplementary Table S4 and Supplementary Table S5).

**Table 2:**
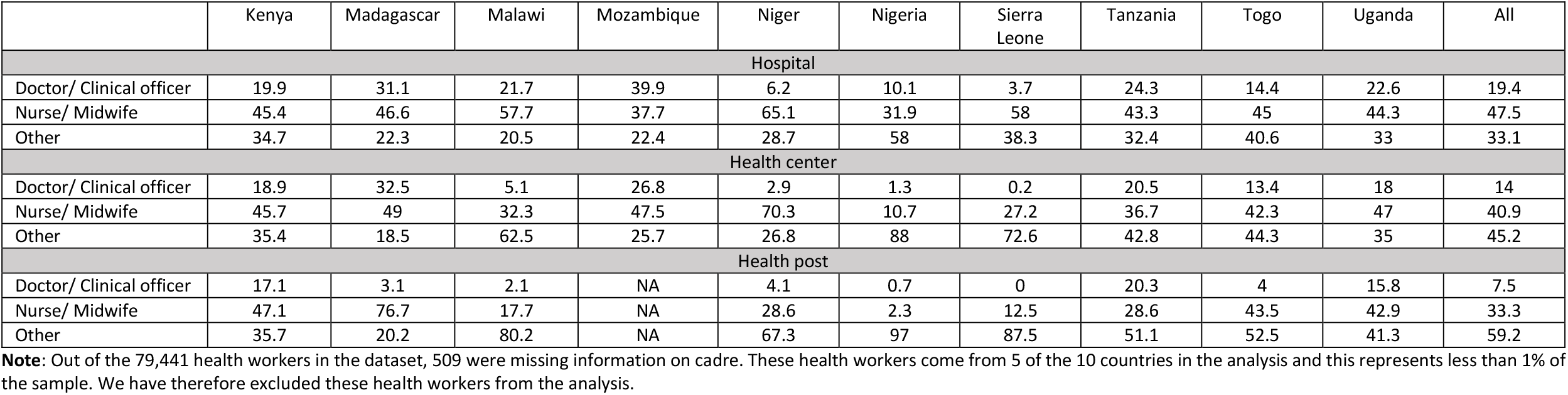
Distribution of health worker cadres by facility type, by country (%)

**Table 3:** Clinical staffing patterns in health centers and health posts, by country (number/proportion and 95% CI)

|  | Kenya | Madagascar | Malawi | Mozambique | Niger | Nigeria | Sierra Leone | Tanzania | Togo | Uganda | All |
| --- | --- | --- | --- | --- | --- | --- | --- | --- | --- | --- | --- |
|  | n = 2753 | n = 407 | n = 1005 | n = 157 | n = 239 | n = 1974 | n = 506 | n = 353 | n = 164 | n = 385 | n = 7943 |
| Average number of clinical staff in health centers and health posts | 3.35<br>(3.15 - 3.55) | 2.17<br>(1.92 - 2.41) | 3.37<br>(3.12 - 3.62) | 5.87<br>(4.33 - 7.4) | 1.38<br>(0.95 - 1.82) | 0.58<br>(0.5 - 0.67) | 1.20<br>(0.95 - 1.44) | 4.36<br>(3.71 - 5) | 2.60<br>(1.88 - 3.33) | 3.42<br>(3.05 - 3.79) | 2.83<br>(1.69 - 3.97) |
| Proportion of health centers and health posts with one or fewer clinical staff | 33.1<br>(31.3 - 35.0) | 50.0<br>(40.6 - 59.4) | 31.9<br>(29.1 - 34.8) | 37.6<br>(30.0 - 45.2) | 79.2<br>(73.1 - 85.3) | 88.1<br>(86.4 - 89.8) | 77.5<br>(73.3 - 81.6) | 22.0<br>(16.1 - 28) | 48.2<br>(32.5 - 63.8) | 28.7<br>(23.2 - 34.3) | 49.6<br>(32.7 - 66.6) |
| Proportion of health centers and health posts with two or fewer clinical staff | 62.1<br>(60.1 - 64) | 82.4<br>(76.8 - 88.1) | 53.6<br>(50.5 - 56.7) | 58.6<br>(50.9 - 66.3) | 88.0<br>(83.5 - 92.4) | 93.5<br>(92.2 - 94.8) | 87.4<br>(83.9 - 90.9) | 47.6<br>(39.9 - 55.4) | 67.0<br>(52.2 - 81.8) | 54.5<br>(48.2 - 60.8) | 69.5<br>(57.5 - 81.5) |

When we examined the distribution of health worker cadres by facility type, we found a wide range of variation by country (Table 2). In some countries such as Kenya and Uganda, health worker cadres showed a similar distribution across facility types with the nurse/midwife being the primary cadre, followed by a substantial proportion of non-clinical/other workers, and supported by a smaller proportion of doctors/clinical officers. In other countries such as Malawi, Niger, and Nigeria, we found health posts were primarily staffed by non-clinical/other health workers, health centers were primarily staffed by nurses/midwives and non-clinical/other health workers, and hospitals were staffed primarily by nurses/midwives supported by doctors/clinical officers and non-clinical/other health workers. Across all countries combined, we found that hospitals had the largest proportion of doctors/clinical officers (19.4% compared to 14% for health centers and 7.5% for health posts) and the largest proportion of nurse/midwives (47.5% compared to 40.9% for health centers and 33.3% for health posts).

When we examined staffing patterns (Table 3), we found that on average health centers and health posts had 2.8 clinical staff, but many facilities had one or no clinical staff. In Madagascar, Niger, Nigeria, and Sierra Leone more than half of health centers and health posts had one or no clinical staff.^11^ In addition, nearly 70% of health centers and health posts across all countries had two or fewer clinical staff. This lack of clinical staff may not be indicative of no patient care provision at these facilities; however, it may suggest that patient care is being provided by under-qualified individuals.^12^

These average levels of clinical staff are far below staffing norms. Supplementary Table S4 shows staffing norms—by cadre and health facility type—for each country. Supplementary Table S5 shows the average staffing norms across cadres and types, compared with the average staffing levels from Table 3. In every case, the staff is less than half that listed in country staffing plans.

### 3.2. Are health workers present? Absenteeism levels, effective staffing patterns, and correlates

We first looked at the overall health worker absenteeism to understand the extent to which health workers who are expected to be present at health facilities are not present for work (Supplementary Table S6). We then examined staffing patterns adjusted for absenteeism to see the impact of absenteeism on the proportion of facilities with limited clinical staff present (Table 4). Finally, we explored the relationship between absenteeism and facility and health worker characteristics to further understand if health workers with certain characteristics were more likely to be absent (Table 5).

**Table 4:**
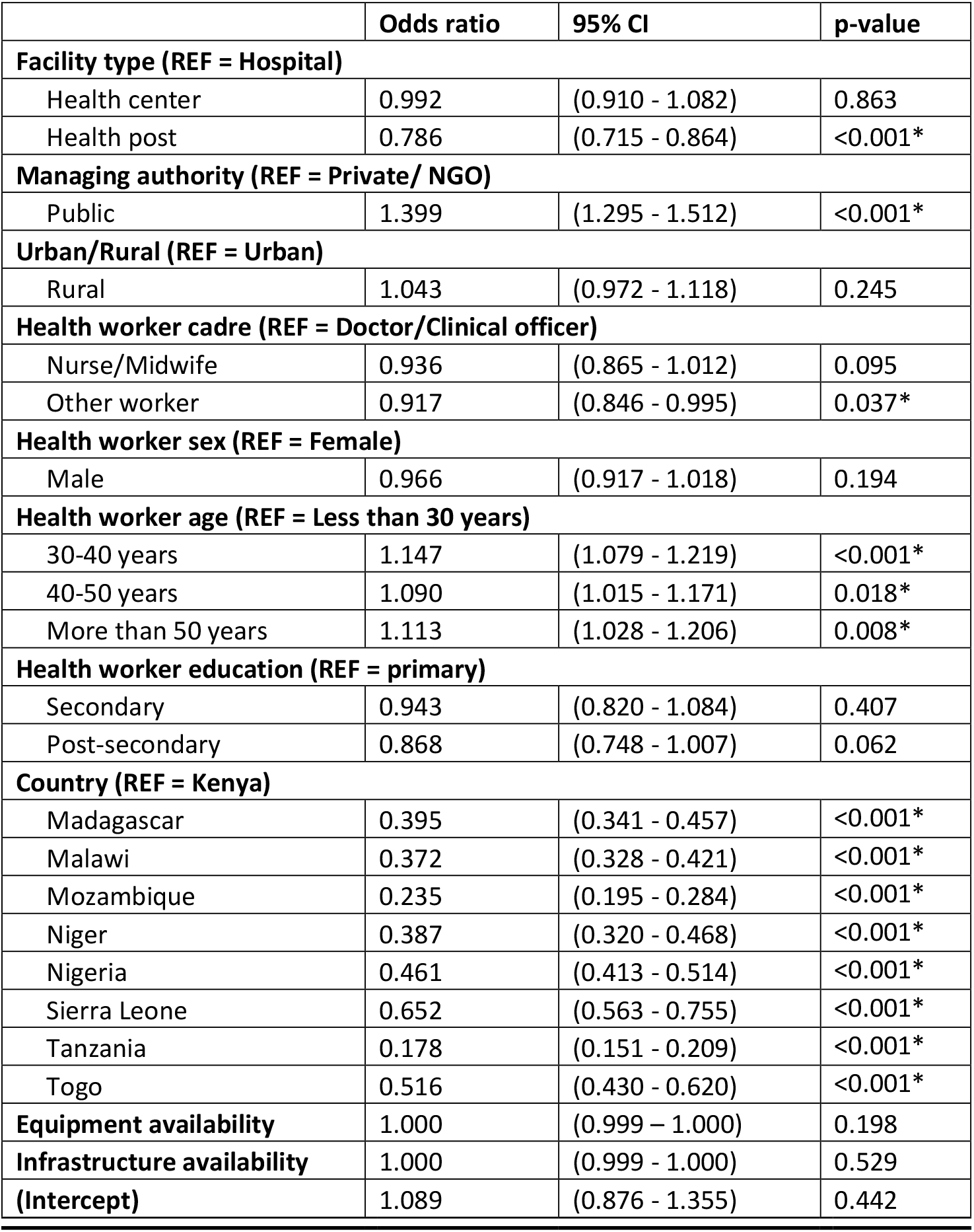
Logistic regression of health worker absenteeism on facility and health worker characteristics.

**Table 5:** Average health worker caseload per facility, by country (number and 95% CI)

|  | Kenya | Madagascar | Mozambique | Niger | Nigeria | Sierra Leone | Tanzania | Togo | Uganda | All |
| --- | --- | --- | --- | --- | --- | --- | --- | --- | --- | --- |
| National | 23.1<br>(21.9 - 24.2) | 6.4<br>(5.0 - 7.7) | 23.2<br>(20.0 - 26.5) | 10.9<br>(8.6 - 13.2) | 2.8<br>(2.5 - 3.2) | 11.3<br>(10.2 - 12.5) | 11.7<br>(9.6 - 13.8) | 6.3<br>(4.8 - 7.9) | 17.0<br>(14.2 - 19.7) | 12.5<br>(6.9 - 18.1) |
| Facility type |  |  |  |  |  |  |  |  |  |  |
| Hospital | 24.6<br>(19.9 - 29.2) | 5.5<br>(3.1 - 7.8) | 16.7<br>(7.9 - 25.5) | 1.1<br>(0.6 - 1.7) | 4.1<br>(2.9 - 5.4) | 8.0<br>(6.3 - 9.8) | 33.5<br>(5.5 - 61.4) | 6.6<br>(4.0 - 9.3) | 16.6<br>(2.9 - 30.2) | 13.0<br>(4.7 - 21.2) |
| Health center | 25.0<br>(22.4 - 27.5) | 7.7<br>(5.5 - 9.9) | 24.9<br>(21.5 - 28.3) | 21.6<br>(17.1 - 26.2) | 2.7<br>(2.3 - 3.0) | 11.3<br>(8.7 - 13.9) | 14.2<br>(7.6 - 20.9) | 7.6<br>(4.5 - 10.6) | 17.1<br>(11.8 - 22.4) | 14.7<br>(8.5 - 20.9) |
| Health post | 22.2<br>(21 - 23.5) | 4.0<br>(2.8 - 5.2) | NA | 6.1<br>(4.5 - 7.6) | 2.0<br>(1.7 - 2.3) | 11.5<br>(10.1 - 12.9) | 10.4<br>(8.5 - 12.4) | 5.8<br>(4.1 - 7.6) | 16.9<br>(13.7 - 20.2) | 9.9<br>(4.1 - 15.6) |
| Managing authority |  |  |  |  |  |  |  |  |  |  |
| Private/NGO | 15.0<br>(13.5 - 16.6) | 11.8<br>(9.8 - 13.7) | 12.5<br>(0.0 - 29.0) | 5.0<br>(2.6 - 7.4) | 3.2<br>(1.9 - 4.6) | 9.2<br>(7.1 - 11.3) | 11.3<br>(7.0 - 15.6) | 6.8<br>(4.3 - 9.4) | 7.0<br>(4.7 - 9.4) | 9.1<br>(6.1 - 12.1) |
| Public | 30.4<br>(28.9 - 32.0) | 5.1<br>(3.6 - 6.6) | 23.4<br>(20.0 - 26.7) | 11.2<br>(8.7 - 13.6) | 2.7<br>(2.5 - 3.0) | 11.5<br>(10.2 - 12.8) | 11.9<br>(9.4 - 14.3) | 6.0<br>(4.0 - 8.0) | 26.1<br>(22.0 - 30.3) | 14.3<br>(6.6 - 21.9) |
| Urban/Rural |  |  |  |  |  |  |  |  |  |  |
| Urban | 20.1<br>(17.5 - 22.6) | 10.5<br>(8.7 - 12.3) | 19.6<br>(5.6 - 33.6) | 8.1<br>(4.5 - 11.7) | 3.2<br>(2.5 - 3.9) | 10.5<br>(8.5 - 12.5) | 15.3<br>(9.7 - 20.9) | 6.6<br>(4.3 - 9.0) | 10.1<br>(5.7 - 14.6) | 11.6<br>(7.2 - 16.0) |
| Rural | 24.6<br>(23.4 - 25.7) | 4.9<br>(3.3 - 6.6) | 23.7<br>(20.5 - 26.9) | 11.2<br>(8.7 - 13.8) | 2.6<br>(2.3 - 2.9) | 11.7<br>(10.2 - 13.2) | 10.1<br>(8.4 - 11.8) | 6.1<br>(4.0 - 8.2) | 20.2<br>(16.8 - 23.5) | 12.8<br>(6.5 - 19.1) |

We found the rate of overall health worker absence was, on average, 34.7% (CI: 26.3% - 43%). On average, across all countries, absenteeism was similarly high across all types of health facilities (hospital 33.6%, health center 35.2%, health post 31.2%), all cadres of health workers (doctors/clinical officers 36.8%, nurses/midwives 34.8%, other workers 31.8%), and in both urban and rural facilities (urban 34.8%, rural 34.6%). Across all countries, on average, a patient was more likely to find a health worker in a private/NGO facility compared to a public facility, but absenteeism was high across facilities of all managing authorities (public 36.4%, private/NGO 29.7%) (Supplementary Table S6, Figure 1).

**Figure 1:**
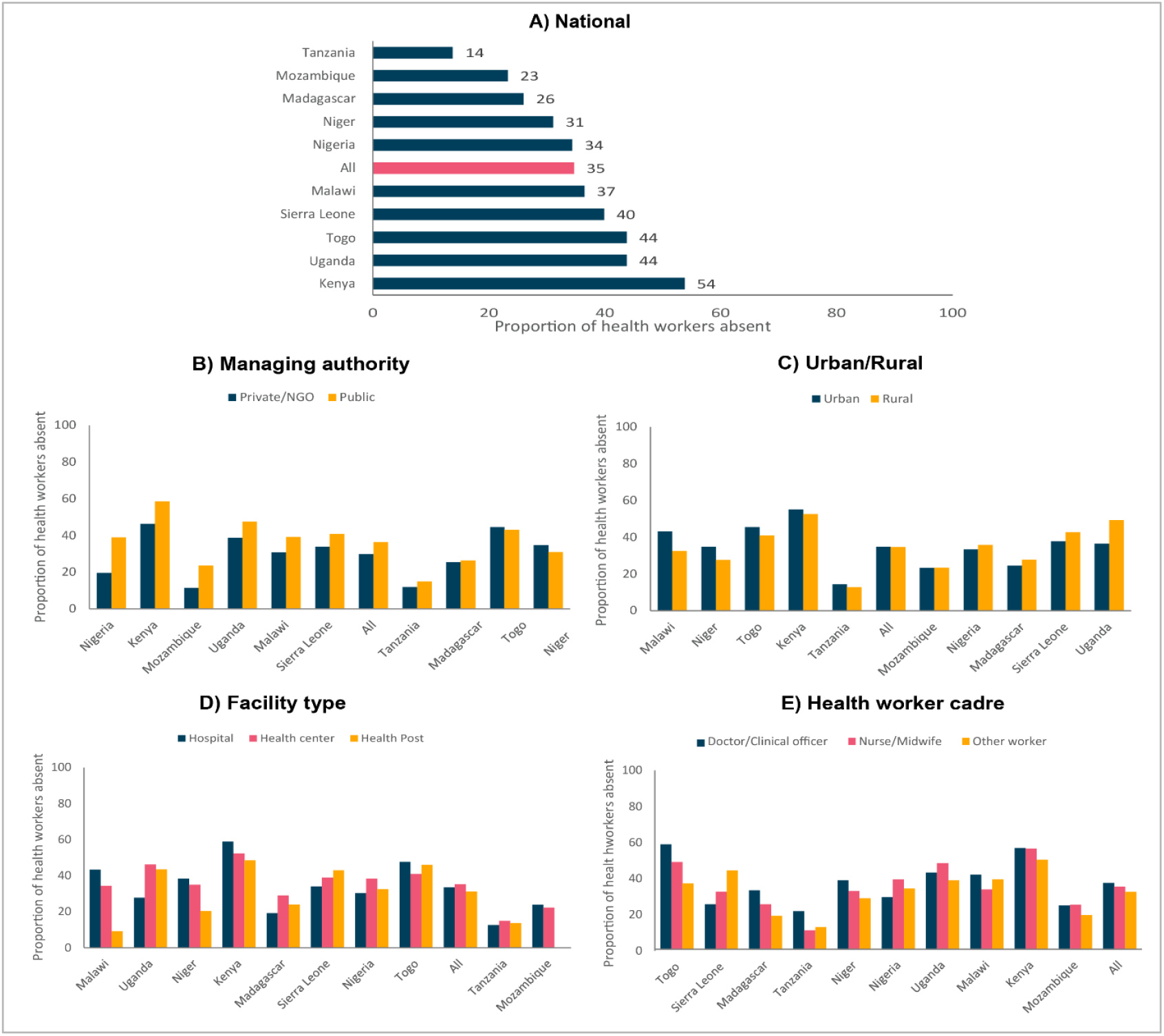
National and sub-national variation in absenteeism

However, there was substantial variation in absenteeism both within and between countries. The proportion of health workers absent from facilities ranged from 53.8% in Kenya to 13.8% in Tanzania. Absenteeism was statistically significantly higher in public facilities than in private facilities in three countries (Nigeria 19 percentage points (pp) higher, Kenya 12 pp higher, and Malawi 9 pp higher). While not statistically significant, five additional countries showed similar trends in higher absenteeism in the public sector, albeit smaller in magnitude. Two countries, Togo and Niger, had diverging findings with absenteeism slightly higher in the private sector (2 and 4 pp respectively), although not statistically significant. While overall, absenteeism was similar in urban and rural areas, this varied by country with about half of countries having higher absenteeism in urban areas and half of countries having higher absenteeism in rural areas. However, only one country, Malawi, had a statistically significant difference between absenteeism in urban and rural facilities with absenteeism being higher in urban facilities (urban 43.1%, rural, 32.5%). No strong pattern emerged across countries related to absenteeism by facility type or health worker cadre. We did not see a single facility type or health worker cadre have consistently higher absenteeism compared to other groups across countries (Supplementary Table S6, Figure 1).

While absenteeism was fairly widespread, further analysis into the reasons for health worker absences revealed that there was very little unauthorized absence. Across all countries, the rate of unauthorized health worker absence was, on average, 3.9% (CI: 1.8% - 5.9%) and no country had an unauthorized health worker absence rate above 9% (Supplementary Table S7). There were a broad range of reasons given for health worker absences including authorized absence, official mission, outreach or fieldwork, sick or maternity leave, training/seminar/meeting, and unauthorized absence (Supplementary Table S8). Our findings indicate that high absenteeism is not indicative of a moral failure on the part of health workers, instead is a characteristic of the health system.

Given the relatively high levels of absenteeism, we examined staffing patterns adjusted for absenteeism to see the impact of absenteeism on the proportion of facilities with limited clinical staff present (Supplementary Table S9, Figure 2). Due to the different distribution of primary and secondary facilities across countries, this analysis focused on primary care facilities only. After adjusting for absenteeism, the staffing patterns in health centers and health posts were much worse off. Across all countries, the proportion of health centers and health posts with one or no clinical staff increased from 49.6% to 64.1% and the proportion of health centers and health posts with two or fewer clinical staff increased from 69.5% to 79.3% after adjusting for absenteeism. In addition, more than half of health centers and health posts in seven out of ten countries had one or no clinical staff after adjusting for absenteeism. The countries in which adjusting for absenteeism had the largest impact on the proportion of health centers and health posts with one or no clinical staff include Kenya (33.1% to 63.9%), Togo (48.2% to 68.9%), and Uganda (28.7% to 59.0%). Adjusting for absenteeism had a much smaller impact on countries where many facilities were already lacking clinical staff before adjusting for absenteeism such as in Niger, Nigeria, and Sierra Leone.

**Figure 2:**
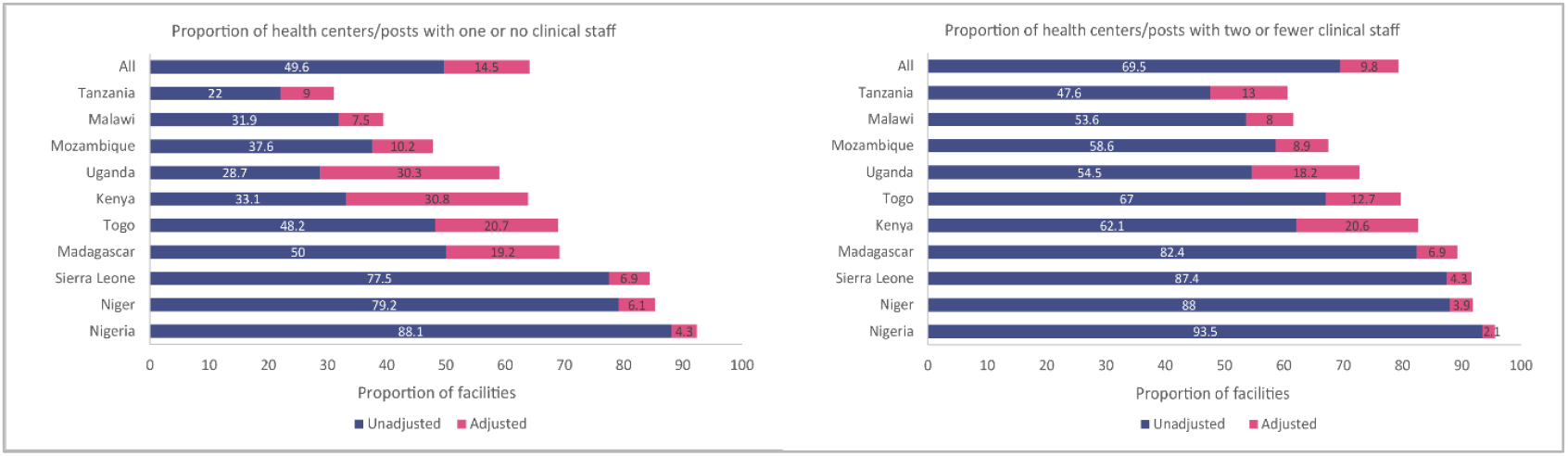
Clinical staffing patterns by country, unadjusted and adjusted for absenteeism at primary care level

Results from the logistic regression of health worker absenteeism on facility and health worker characteristics indicate that both facility and health worker level characteristics are associated with absenteeism (Table 4). The facility level covariates that were statistically significantly associated with absenteeism include facility type and managing authority. Absenteeism was higher for health workers at public facilities (β = 1.399, p-value <0.001) compared to private facilities and lower for health workers at health posts (β = 0.786, p-value <0.001) as compared to health workers at hospitals. Interestingly, there was no difference in absenteeism between rural and urban health workers after controlling for other facility and health worker characteristics. The health worker level covariates that were statistically significantly associated with absenteeism include cadre and age. Absenteeism was lower for other workers (β = 0.917, p-value = 0.037) as compared to doctors while absenteeism was higher for health workers of all age groups compared to health workers less than 30 years. Our findings demonstrate that absenteeism was lowest at the smallest health facilities while being substantially higher at public facilities and slightly higher for older health workers.

### 3.3. How heavy are health workers’ caseloads?

In order to understand staffing needs beyond the proportion of facilities with only one or two clinical staff, we first looked at health worker caseload to understand how many clients health workers are seeing per day (Table 5). We then examined the proportion of facilities with low and high caseload to understand how many facilities are potentially not maximizing productivity (Table 6). Finally, we explored the relationship between caseload and facility characteristics to further understand if facilities with certain characteristics were more likely to have higher caseloads (Table 7). Malawi was excluded from the caseload analysis due to potential discrepancies in the collection of data on outpatient visits. The caseload analysis including Malawi is provided in the supplementary materials (Supplementary Tables S10, S11, S12 and Supplementary Figures S1a and S1b).

**Table 6:** Proportion of facilities with low and high caseload, by country (% and 95% CI)

|  | Kenya | Madagascar | Mozambique | Niger | Nigeria | Sierra Leone | Tanzania | Togo | Uganda | All |
| --- | --- | --- | --- | --- | --- | --- | --- | --- | --- | --- |
| **Proportion of facilities with a caseload of <5** |  |  |  |  |  |  |  |  |  |  |
| Private/NGO | 31.7<br>(28.7 - 34.6) | 30.7<br>(19.7 - 41.8) | 50.0<br>(0.0 - 119.9) | 70.5<br>(52.6 - 88.3) | 83.1<br>(76.2 - 89.9) | 32.5<br>(16.8 - 48.1) | 40.5<br>(24.7 - 56.4) | 53.2<br>(23.3 - 83.2) | 64.1<br>(54.1 - 74.2) | 50.7<br>(36.3 - 65.1) |
| Public | 5.0<br>(3.8 - 6.1) | 73.1<br>(64.8 - 81.5) | 13.6<br>(8.6 - 18.6) | 40.7<br>(30.3 - 51.1) | 86.5<br>(84.7 - 88.3) | 32.3<br>(27.6 - 37.1) | 35.6<br>(26.7 - 44.5) | 63.9<br>(50.2 - 77.7) | 3.4<br>(0.5 - 6.3) | 39.3<br>(16.3 - 62.3) |
| Total | 17.8<br>(16.1 - 19.4) | 65.2<br>(57.0 - 73.4) | 14.0<br>(9.0 - 19.0) | 41.8<br>(31.8 - 51.8) | 85.9<br>(84.0 - 87.8) | 32.4<br>(27.8 - 36.9) | 37.0<br>(29.2 - 44.7) | 59.5<br>(44.9 - 74.1) | 32.5<br>(25.6 - 39.4) | 42.9<br>(25.0 - 60.8) |
| **Proportion of facilities with a caseload of >=30** |  |  |  |  |  |  |  |  |  |  |
| Private/NGO | 12.8<br>(10.5 - 15.1) | 9.1<br>(4.3 - 13.9) | 0.0<br>(0.0 - 0.0) | 0.0<br>(0.0 - 0.0) | 1.9<br>(0.0 - 4.4) | 0.0<br>(0.0 - 0.0) | 3.7<br>(0.1 - 7.3) | 0.0<br>(0.0 - 0.0) | 4.7<br>(0.7 - 8.7) | 3.6<br>(0.0 - 7.1) |
| Public | 36.1<br>(33.4 - 38.7) | 1.2<br>(0.0 - 2.3) | 27.2<br>(20.7 - 33.7) | 10.7<br>(4.1 - 17.2) | 0.8<br>(0.3 - 1.3) | 6.2<br>(4.1 - 8.4) | 6.5<br>(3.8 - 9.2) | 3.8<br>(0.0 - 11.2) | 29.8<br>(22.4 - 37.2) | 13.6<br>(3.1 - 24.0) |
| Total | 24.9<br>(23.1 - 26.8) | 2.6<br>(1.3 - 4.0) | 26.9<br>(20.5 - 33.3) | 10.3<br>(4.0 - 16.5) | 1.0<br>(0.4 - 1.6) | 5.7<br>(3.7 - 7.7) | 5.7<br>(3.5 - 7.9) | 2.2<br>(0.0 - 6.6) | 17.8<br>(13.2 - 22.3) | 10.8<br>(3.1 - 18.5) |

**Table 7:**
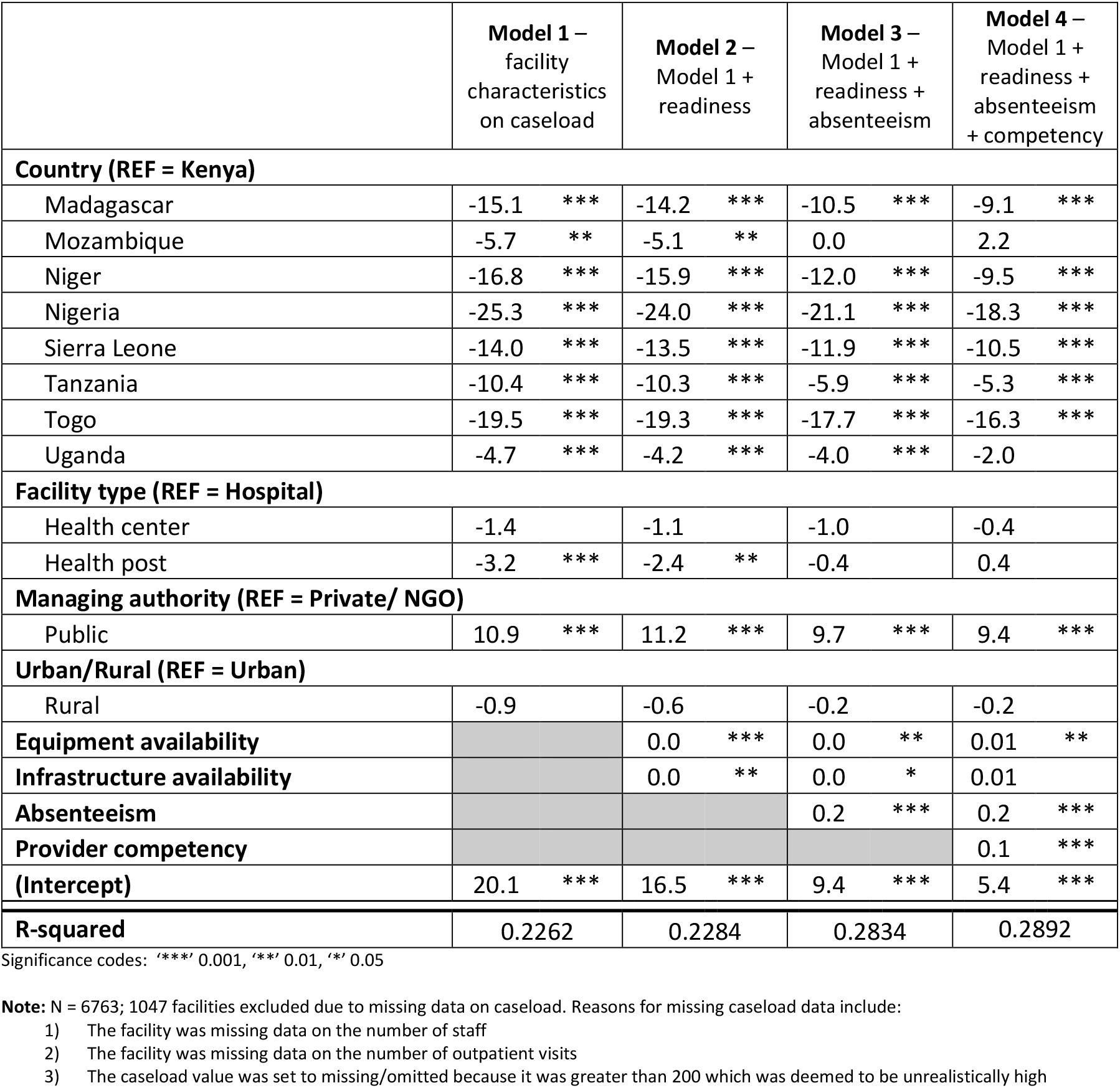
Linear regression of caseload on facility characteristics.

|  | **Model 1 –**<br>**facility**<br>**characteristics**<br>**on caseload** | **Model 2 –**<br>**Model 1 +**<br>**readiness** | **Model 3 –**<br>**Model 1 +**<br>**readiness +**<br>**absenteeism** | **Model 4 –**<br>**Model 1 +**<br>**readiness +**<br>**absenteeism**<br>**+ competency** |
| --- | --- | --- | --- | --- |
| **Country (REF = Kenya)** |  |  |  |  |
| Madagascar | -15.1 *** | -14.2 *** | -10.5 *** | -9.1 *** |
| Mozambique | -5.7 ** | -5.1 ** | 0.0 | 2.2 |
| Niger | -16.8 *** | -15.9 *** | -12.0 *** | -9.5 *** |
| Nigeria | -25.3 *** | -24.0 *** | -21.1 *** | -18.3 *** |
| Sierra Leone | -14.0 *** | -13.5 *** | -11.9 *** | -10.5 *** |
| Tanzania | -10.4 *** | -10.3 *** | -5.9 *** | -5.3 *** |
| Togo | -19.5 *** | -19.3 *** | -17.7 *** | -16.3 *** |
| Uganda | -4.7 *** | -4.2 *** | -4.0 *** | -2.0 |
| **Facility type (REF = Hospital)** |  |  |  |  |
| Health center | -1.4 | -1.1 | -1.0 | -0.4 |
| Health post | -3.2 *** | -2.4 ** | -0.4 | 0.4 |
| **Managing authority (REF = Private/ NGO)** |  |  |  |  |
| Public | 10.9 *** | 11.2 *** | 9.7 *** | 9.4 *** |
| **Urban/Rural (REF = Urban)** |  |  |  |  |
| Rural | -0.9 | -0.6 | -0.2 | -0.2 |
| **Equipment availability** |  | 0.0 *** | 0.0 ** | 0.01 ** |
| **Infrastructure availability** |  | 0.0 ** | 0.0 * | 0.01 |
| **Absenteeism** |  |  | 0.2 *** | 0.2 *** |
| **Provider competency** |  |  |  | 0.1 *** |
| **(Intercept)** | 20.1 *** | 16.5 *** | 9.4 *** | 5.4 *** |
| **R-squared** | 0.2262 | 0.2284 | 0.2834 | 0.2892 |

Significance codes: '\*\*\*' 0.001, '\*\*' 0.01, '\*' 0.05
**Note:** N = 6763; 1047 facilities excluded due to missing data on caseload. Reasons for missing caseload data include:
- 1) The facility was missing data on the number of staff - 2) The facility was missing data on the number of outpatient visits - 3) The caseload value was set to missing/omitted because it was greater than 200 which was deemed to be unrealistically high

When examining health worker caseload, we found that on average a health worker attended to 12.5 outpatients per day (CI: 6.9 – 18.1). On average, across all countries, caseload was similar across all types of health facilities (hospital 13, health center 14.7, health post 9.9), in both private/NGO and public facilities (private/NGO 9.1, public 14.3) and in both urban and rural facilities (urban 11.6, rural 12.8). However, health worker caseload varied substantially both across and within countries. Health worker caseload ranged from a low of 2.8 outpatients per day (CI: 2.5 - 3.2) in Nigeria to a high of 23.2 outpatients per day in Mozambique (CI: 20 – 26.5) and 23.1 outpatients per day in Kenya (CI: 21.9 – 24.2). While there is no global benchmark for caseload, some basic assumptions can be made to better contextualize caseload values. Assuming a health worker saw one patient every half hour over the course of an eight-hour shift, this would be a caseload of 16 outpatients per day. Six out of nine countries (Madagascar, Niger, Nigeria, Sierra Leone, Tanzania, and Togo) had an average caseload lower than 16 outpatients per day. Caseload was statistically significantly higher in public facilities than in private facilities in three countries (Kenya 15 pp higher, Niger 6 pp higher, and Uganda 19 pp higher) while one country, Madagascar, had diverging findings with caseload statistically significantly higher in the private sector (7 pp). While overall caseload was similar in urban and rural areas, this varied by country with about half of countries demonstrating higher caseload in urban areas and half of countries demonstrating higher caseload in rural areas. However, only three countries had a statistically significant difference between caseload in urban and rural facilities with caseload being higher in urban facilities in Madagascar (urban 10.5, rural, 4.9) and higher in rural facilities in Kenya (urban 20.1, rural, 24.6) and Uganda (urban 10.1, rural, 20.2). No strong pattern emerged across countries related to caseload by facility type. We did not see a single facility type have consistently higher caseload compared to other groups across countries (Table 5).

We then examined the proportion of facilities with very low and very high caseload, both which may be suggestive of health system inefficiencies. The distribution of caseload by country shows that in countries such as Mozambique, Kenya, and Uganda there were some facilities with a much higher caseload than other facilities while in other countries such as Nigeria, Togo, and Madagascar, caseload was quite similar across all facilities. The high inter-quartile range (IQR) for caseload in Mozambique (23.2), Kenya (22.9), and Uganda (19.0) is indicative of substantial within country variation in caseload. Conversely, the low IQR for caseload in Nigeria (2.2), Togo (4.5), and Madagascar (4.7) is indicative of little within country variation in caseload. The distribution of caseload also points to several countries in which most facilities had a rather low caseload (Figure 3). We assessed the proportion of facilities with a caseload of less than 5 outpatients per day (low caseload) and the proportion of facilities with a caseload greater than or equal to 30 outpatients per day (high caseload). Across all countries, 42.9% of facilities had a caseload of less than 5 outpatients per day. The proportion of facilities with a caseload of fewer than 5 outpatients per day ranged from 14.0% in Mozambique to 85.9% in Nigeria. More than half of facilities in Madagascar, Nigeria, and Togo had a caseload of less than 5 outpatients per day. In three countries (Kenya, Niger, and Uganda), the proportion of facilities with a caseload of less than 5 outpatients per day was substantially higher for the private/NGO sector as compared to the public sector while in one country, Madagascar, the proportion of facilities with a caseload of less than 5 outpatients per day was substantially higher for the public sector as compared to the private sector. Across all countries, 10.8% of facilities had a caseload of 30 or more outpatients per day. The proportion of facilities with a caseload of 30 or more outpatients per day ranged from 1.0% in Nigeria to 26.9% in Mozambique. Approximately one-quarter of facilities in Kenya and Mozambique had a caseload of 30 or more outpatients per day while 10% or less of facilities in Madagascar, Niger, Nigeria, Tanzania, and Togo had a caseload of 30 or more. In five countries (Kenya, Mozambique, Niger, Sierra Leone, and Uganda), the proportion of facilities with a caseload of 30 or more outpatients per day was substantially higher for the public sector as compared to the private (Table 6).

**Figure 3:**
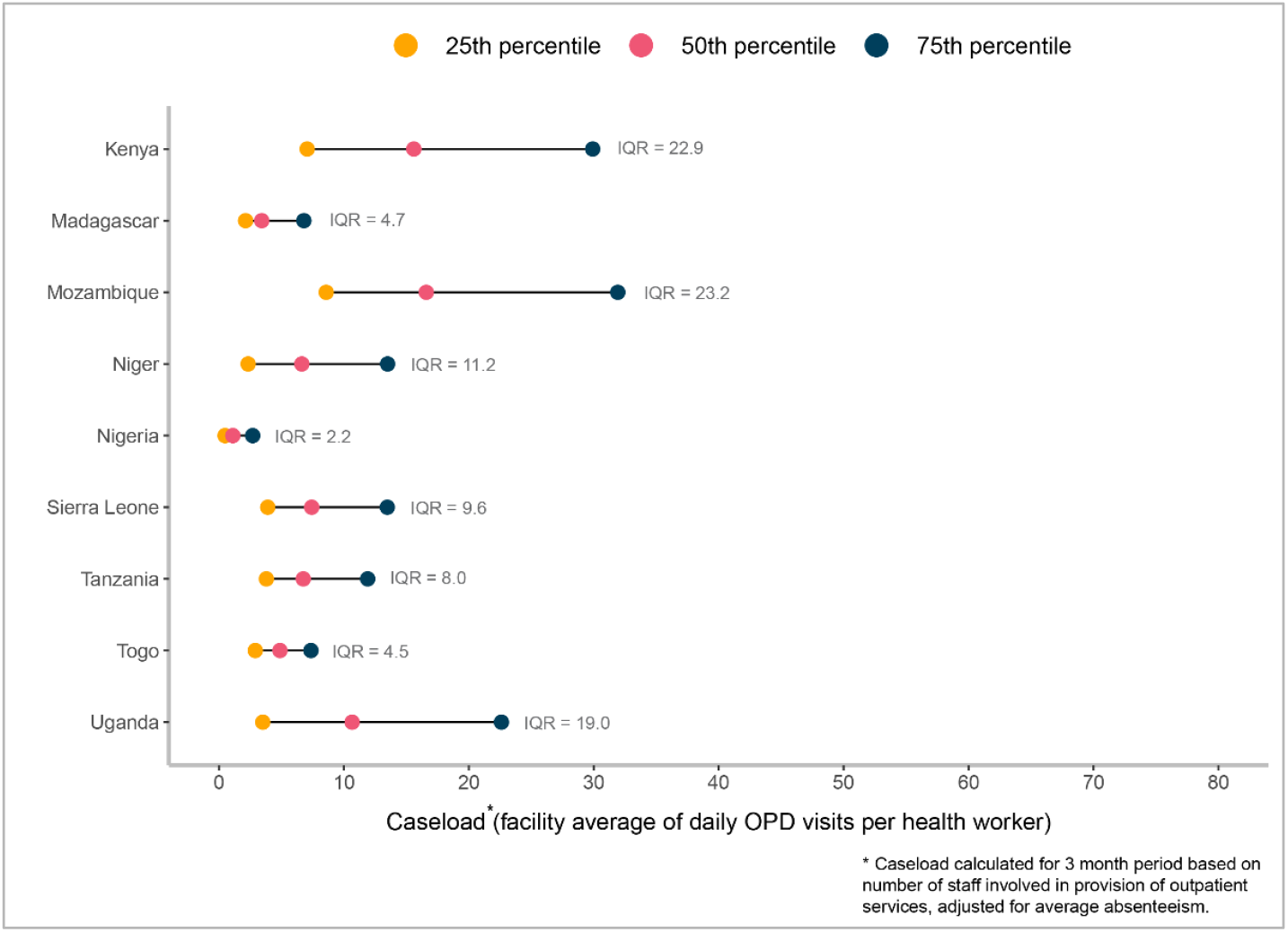
Caseload distribution, by country

Finally, we explored associations between caseload and facility characteristics to further understand if facilities with certain characteristics were more likely to have higher or lower caseloads. We first examined the relationship between caseload and basic facility characteristics and then included additional explanatory variables such as readiness, absenteeism, and provider competency.^13^ Results from the linear regression of caseload on facility characteristics, readiness, absenteeism, and competency indicate that public facilities have much higher caseloads than private/NGO facilities (β = 9.371, p-value <0.001). In addition, caseload is modestly higher when both absenteeism (β = 0.189, p-value <0.001) and competency are higher (β = 0.073, p-value <0.001). There was no difference in caseload between urban and rural facilities, nor between health centers and hospitals after controlling for other facility characteristics. In addition, increases in readiness were not all associated with increased caseload, with only increases in equipment availability being associated with a small increase in caseload (β = 0.011, p-value <0.01) (Table 7).

### 3.4. How competent are health workers?

To understand health worker competency, we first looked at a measure of diagnostic and treatment accuracy. As diagnostic accuracy and treatment accuracy are highly correlated, we present here (Table 8) the combined indicator for both diagnostic and treatment accuracy. Detailed results for diagnostic accuracy and treatment accuracy individually can be found in Supplementary Table S13 and Supplementary Table S14, respectively. We then examined the proportion of health workers with a diagnostic and treatment accuracy score of 50% or less, 20% or less, and equal to 0% (Table 9). Finally, we explored the relationship between diagnostic and treatment accuracy and facility and health worker characteristics (Table 10).

**Table 8:**
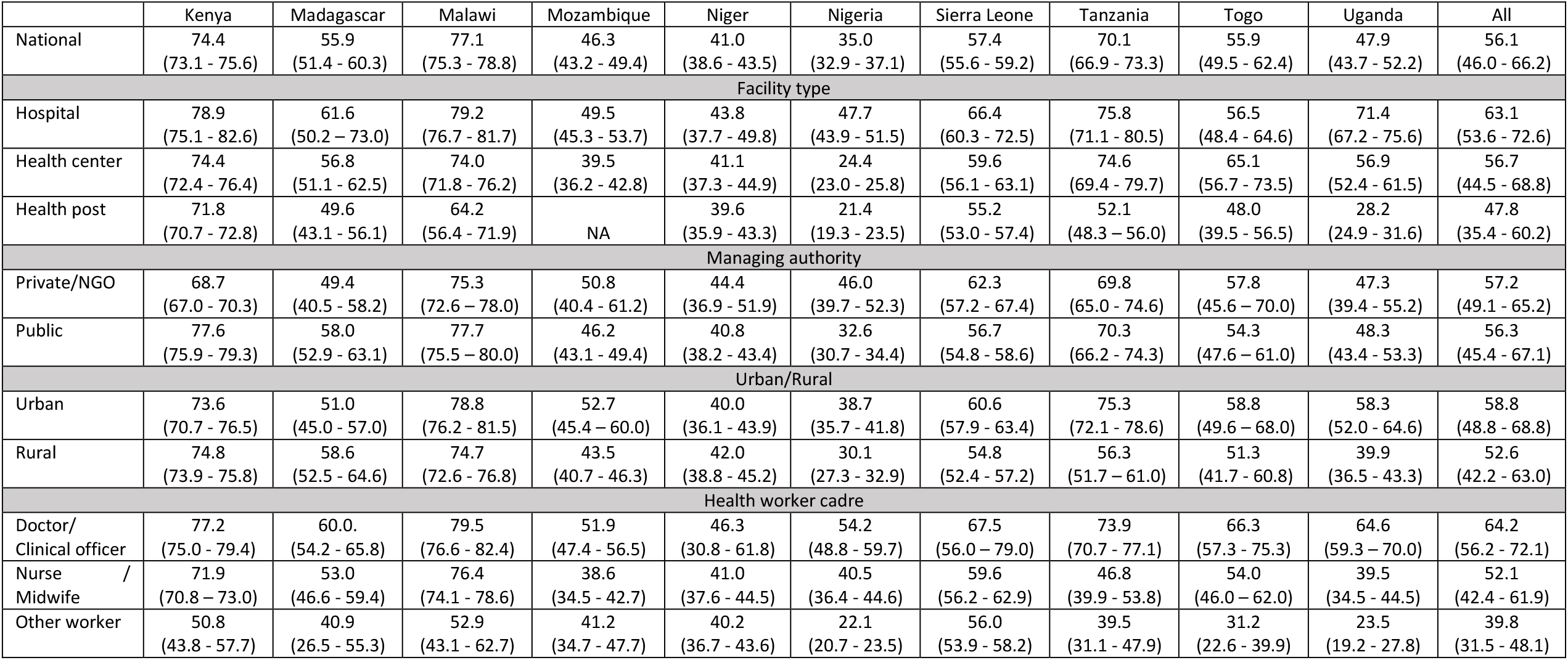
Health worker diagnostic and treatment accuracy, by country (% and 95% CI)

**Table 9:** Proportion of health workers with low competency, by country (% and 95% CI)

|  | Kenya | Madagascar | Malawi | Mozambique | Niger | Nigeria | Sierra Leone | Tanzania | Togo | Uganda | All |
| --- | --- | --- | --- | --- | --- | --- | --- | --- | --- | --- | --- |
| **A. PROPORTION OF HEALTH WORKERS WITH DIAGNOSTIC AND TREATMENT ACCURACY 50% OR LESS** |  |  |  |  |  |  |  |  |  |  |  |
| National | 57.0<br>(53.9 - 60.1) | 89.3<br>(84.4 - 94.2) | 27.7<br>(22.4 - 33.0) | 74.1<br>(68.2 - 80.0) | 91.9<br>(88.9 - 94.9) | 85.8<br>(82.7 - 88.9) | 76.8<br>(73.5 - 80.2) | 30.2<br>(20.5 - 40.0) | 70.4<br>(58.1 - 82.8) | 75.8<br>(67.2 - 84.3) | 67.9<br>(51.6 - 84.2) |
| Health worker cadre |  |  |  |  |  |  |  |  |  |  |  |
| Doctor/ Clinical officer | 47.2<br>(42.1 - 52.3) | 85.4<br>(77.7 - 93.1) | 20.4<br>(10.7 - 30.1) | 64.1<br>(54.9 - 73.4) | 75.6<br>(57.7 - 93.6) | 70.5<br>(61.0 - 79.9) | 52.4<br>(30.2 - 74.5) | 22.8<br>(12.3 - 33.3) | 41.0<br>(20.9 - 61.1) | 58.8<br>(40.7 - 77) | 53.8<br>(38.4 - 69.2) |
| Nurse / Midwife | 67.4<br>(65.2 - 69.6) | 92.1<br>(85.7 - 98.6) | 32.0<br>(26.5 - 37.6) | 87.7<br>(82.1 - 93.3) | 92.2<br>(88.0 - 96.5) | 85.9<br>(80.7 - 91.1) | 76.5<br>(70.5 - 82.5) | 80.9<br>(70.9 - 90.8) | 79.0<br>(66.1 - 92.0) | 85.2<br>(75.9 - 94.5) | 77.9<br>(65.2 - 90.6) |
| Other worker | 83.4<br>(74.1 - 92.8) | 100.0<br>(100.0 - 100.0) | 58.0<br>(41.3 - 74.7) | 83.5<br>(72.7 - 94.2) | 94.1<br>(90.0 - 98.2) | 94.4<br>(92.8 - 95.9) | 77.7<br>(73.6 - 81.9) | 80.8<br>(68.0 - 93.6) | 93.7<br>(81.2 - 100.0) | 98.7<br>(96.8 - 100.0) | 86.4<br>(77.4 - 95.5) |
| **B. PROPORTION OF HEALTH WORKERS WITH DIAGNOSTIC AND TREATMENT ACCURACY 20% OR LESS** |  |  |  |  |  |  |  |  |  |  |  |
| National | 6.3<br>(4.2 - 8.4) | 24.2<br>(16.2 - 32.2) | 7.8<br>(5.8 - 9.8) | 44.3<br>(38.4 - 50.2) | 71.1<br>(65.7 - 76.6) | 66.6<br>(62.8 - 70.5) | 49.6<br>(45.5 - 53.7) | 13.3<br>(8.3 - 18.4) | 32.9<br>(22.2 - 43.6) | 42.5<br>(34.5 - 50.5) | 35.9<br>(19.3 - 52.4) |
| Health worker cadre |  |  |  |  |  |  |  |  |  |  |  |
| Doctor/ Clinical officer | 5.2<br>(1.3 - 9.0) | 10.7<br>(4.9 - 16.5) | 2.6<br>(1.0 - 4.2) | 34.3<br>(25.9 - 42.8) | 32.6<br>(8.9 - 56.3) | 33.6<br>(23.2 - 43.9) | 36.0<br>(11.2 - 60.8) | 6.9<br>(2.3 - 11.5) | 29.3<br>(12.1 - 46.6) | 11.9<br>(5.7 - 18.0) | 20.3<br>(10.4 - 30.3) |
| Nurse / Midwife | 6.8<br>(5.6 - 8.1) | 34.6<br>(21.8 - 47.3) | 10.4<br>(7.3 - 13.5) | 57.1<br>(48.9 - 65.4) | 68.1<br>(60.1 - 76.1) | 64.0<br>(57.9 - 70.2) | 41.6<br>(34.3 - 48.8) | 53.4<br>(38.3 - 68.6) | 32.5<br>(18.9 - 46.1) | 59.9<br>(51.1 - 68.6) | 42.8<br>(27.4 - 58.3) |
| Other worker | 31.2<br>(21.3 - 41.2) | 48.1<br>(18.7 - 77.5) | 38.2<br>(23.5 - 53.0) | 55.3<br>(40.3 - 70.2) | 81.6<br>(75.0 - 88.1) | 86.2<br>(84.1 - 88.2) | 53.6<br>(48.6 - 58.6) | 63.9<br>(48.3 - 79.6) | 56.0<br>(17.6 - 94.4) | 82.8<br>(74.1 - 91.5) | 59.7<br>(46.2 - 73.2) |
| **C. PROPORTION OF HEALTH WORKERS WITH DIAGNOSTIC AND TREATMENT ACCURACY EQUAL TO 0%** |  |  |  |  |  |  |  |  |  |  |  |
| National | 6.3<br>(4.2 - 8.4) | 24.2<br>(16.2 - 32.2) | 2.7<br>(1.8 - 3.7) | 14.4<br>(10.4 - 18.4) | 39.1<br>(33.1 - 45.2) | 46.7<br>(43.3 - 50.2) | 22.4<br>(18.9 - 26.0) | 4.8<br>(2.8 - 6.9) | 13.5<br>(7.3 - 19.8) | 24.8<br>(19.5 - 30.1) | 19.9<br>(9.5 - 30.4) |
| Health worker cadre |  |  |  |  |  |  |  |  |  |  |  |
| Doctor/ Clinical officer | 5.2<br>(1.3 - 9) | 10.7<br>(4.9 - 16.5) | 0.4<br>(0.1 - 0.7) | 9.1<br>(3.6 - 14.6) | 24.3<br>(0.1 - 48.6) | 19.9<br>(10 - 29.7) | 5.5<br>(0 - 13.8) | 1.5<br>(0.5 - 2.5) | 12.3<br>(0.1 - 24.4) | 4.1<br>(1.1 - 7.1) | 9.3<br>(3.7 - 14.9) |
| Nurse / Midwife | 6.8<br>(5.6 - 8.1) | 34.6<br>(21.8 - 47.3) | 2.9<br>(1.4 - 4.4) | 22.0<br>(14.9 - 29.1) | 30.4<br>(22.8 - 38.0) | 36.8<br>(31.4 - 42.2) | 17.5<br>(11.8 - 23.2) | 27.1<br>(13.0 - 41.1) | 12.6<br>(5.1 - 20.1) | 32.5<br>(25.5 - 39.5) | 22.3<br>(13.8 - 30.9) |
| Other worker | 31.2<br>(21.3 - 41.2) | 48.1<br>(18.7 - 77.5) | 33.0<br>(19.2 - 46.8) | 18.8<br>(9.3 - 28.2) | 53.6<br>(44.3 - 62.9) | 65.5<br>(63.2 - 67.9) | 25.2<br>(20.7 - 29.7) | 27.8<br>(12.7 - 42.8) | 32.6<br>(5.7 - 59.6) | 62.6<br>(52.8 - 72.3) | 39.8<br>(28.1 - 51.5) |

**Table 10:**
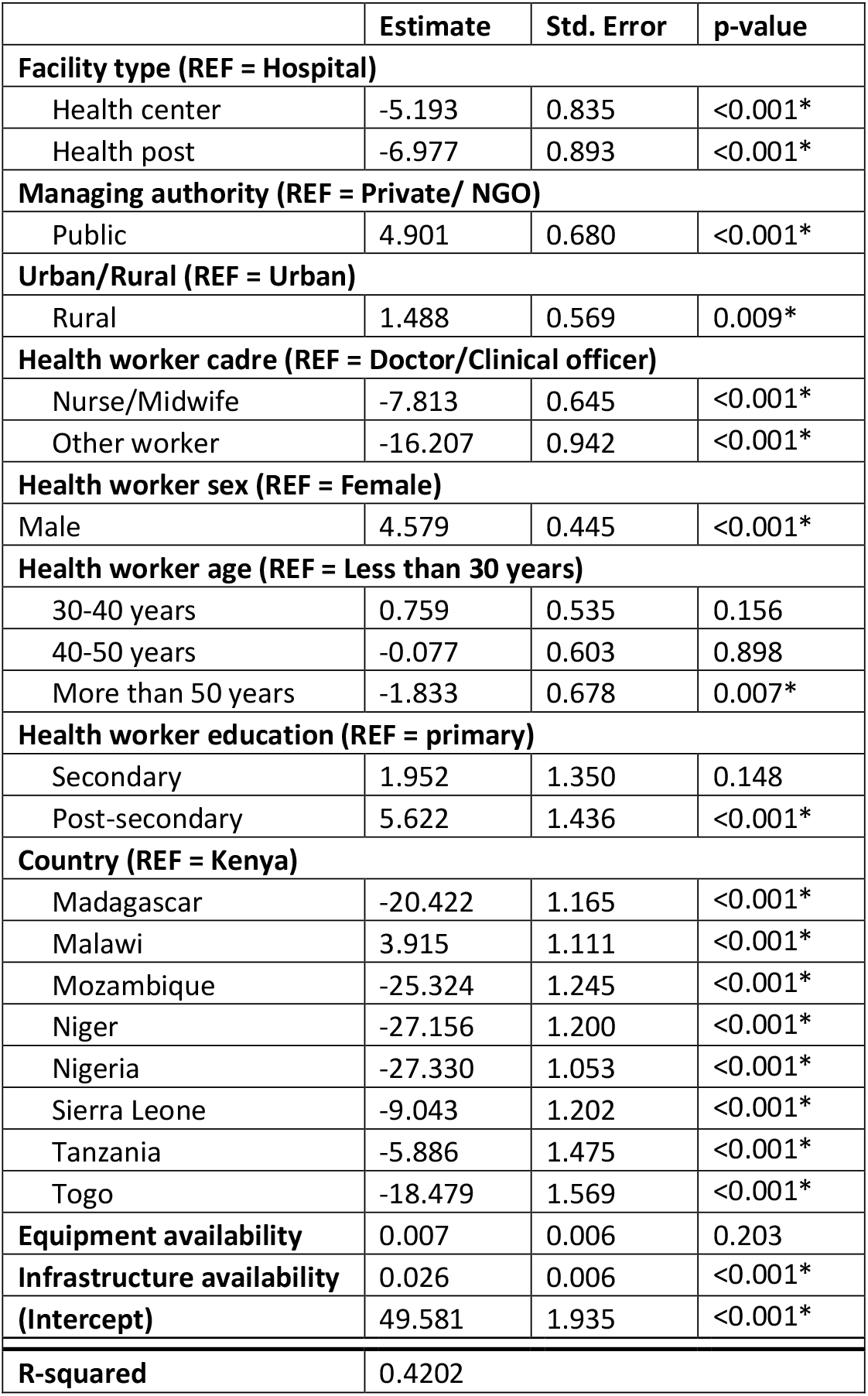
Linear regression of diagnostic and treatment accuracy on facility and health worker characteristics.

|  | Estimate | Std. Error | p-value |
| --- | --- | --- | --- |
| **Facility type (REF = Hospital)** |  |  |  |
| Health center | -5.193 | 0.835 | <0.001* |
| Health post | -6.977 | 0.893 | <0.001* |
| **Managing authority (REF = Private/ NGO)** |  |  |  |
| Public | 4.901 | 0.680 | <0.001* |
| **Urban/Rural (REF = Urban)** |  |  |  |
| Rural | 1.488 | 0.569 | 0.009* |
| **Health worker cadre (REF = Doctor/Clinical officer)** |  |  |  |
| Nurse/Midwife | -7.813 | 0.645 | <0.001* |
| Other worker | -16.207 | 0.942 | <0.001* |
| **Health worker sex (REF = Female)** |  |  |  |
| Male | 4.579 | 0.445 | <0.001* |
| **Health worker age (REF = Less than 30 years)** |  |  |  |
| 30-40 years | 0.759 | 0.535 | 0.156 |
| 40-50 years | -0.077 | 0.603 | 0.898 |
| More than 50 years | -1.833 | 0.678 | 0.007* |
| **Health worker education (REF = primary)** |  |  |  |
| Secondary | 1.952 | 1.350 | 0.148 |
| Post-secondary | 5.622 | 1.436 | <0.001* |
| **Country (REF = Kenya)** |  |  |  |
| Madagascar | -20.422 | 1.165 | <0.001* |
| Malawi | 3.915 | 1.111 | <0.001* |
| Mozambique | -25.324 | 1.245 | <0.001* |
| Niger | -27.156 | 1.200 | <0.001* |
| Nigeria | -27.330 | 1.053 | <0.001* |
| Sierra Leone | -9.043 | 1.202 | <0.001* |
| Tanzania | -5.886 | 1.475 | <0.001* |
| Togo | -18.479 | 1.569 | <0.001* |
| **Equipment availability** | 0.007 | 0.006 | 0.203 |
| **Infrastructure availability** | 0.026 | 0.006 | <0.001* |
| **(Intercept)** | 49.581 | 1.935 | <0.001* |
| **R-squared** | 0.4202 |  |  |

When examining health worker competency, we found diagnostic and treatment accuracy was, on average, 56.1% (CI: 46% - 66.2%), meaning that on average, health workers gave the correct diagnosis and treatment for just over half of cases. On average, across all countries, diagnostic and treatment accuracy was similar in both urban and rural facilities (58.8% and 52.6%) and public and private/NGO facilities (56.3% and 57.2%). However, it differed by facility type and health worker cadre and a decreasing trend in health worker competency was seen as the facility level decreased from hospital (63.1%) to health center (56.7%) to health post (47.8%) and as the health worker level of training decreased from doctor/clinical officer (64.2%) to nurse/midwife (52.1%) to other workers (39.8%).

Diagnostic and treatment accuracy varied by country and was highest in Malawi (77.1%, CI: 75.3% - 78.8%) and lowest in Nigeria (35%, CI: 32.9% - 37.1%). The range of health worker competency between health worker cadres varied substantially by country. The difference in diagnostic and treatment accuracy between doctors/clinical officers and nurses/midwifes ranged from a 27-pp difference in Tanzania to a 3-pp difference in Malawi while the difference in diagnostic and treatment accuracy between nurses/midwives and other workers ranged from a 24-pp difference in Malawi to a 1-pp difference in Niger (Table 8).

We also examined the proportion of health workers with a diagnostic and treatment accuracy score of 50% or less, 20% or less, and equal to 0%. Across all countries, 67.9% of health workers (CI: 51.6% - 84.2%) had a diagnostic and treatment accuracy score of 50% or less, 35.9% of health workers (CI: 19.3% - 52.4%) had a diagnostic and treatment accuracy score of 20% or less, and 19.9% of health workers (CI: 9.5% - 30.4%) had a diagnostic and treatment accuracy score equal to 0%. The proportion of health workers with a competency score of 0, was on average, 9.3% for doctors/clinical officers, 22.3% for nurses/midwives, and 39.8% for other workers (Table 9). However, the proportion of health workers with a competency score of 0 varied by country, with countries such as Madagascar, Niger, and Nigeria, having more than 10% of doctors/clinical officers and more than 30% of nurses/midwives with a competency score of 0 (Figure 4).

**Figure 4:**
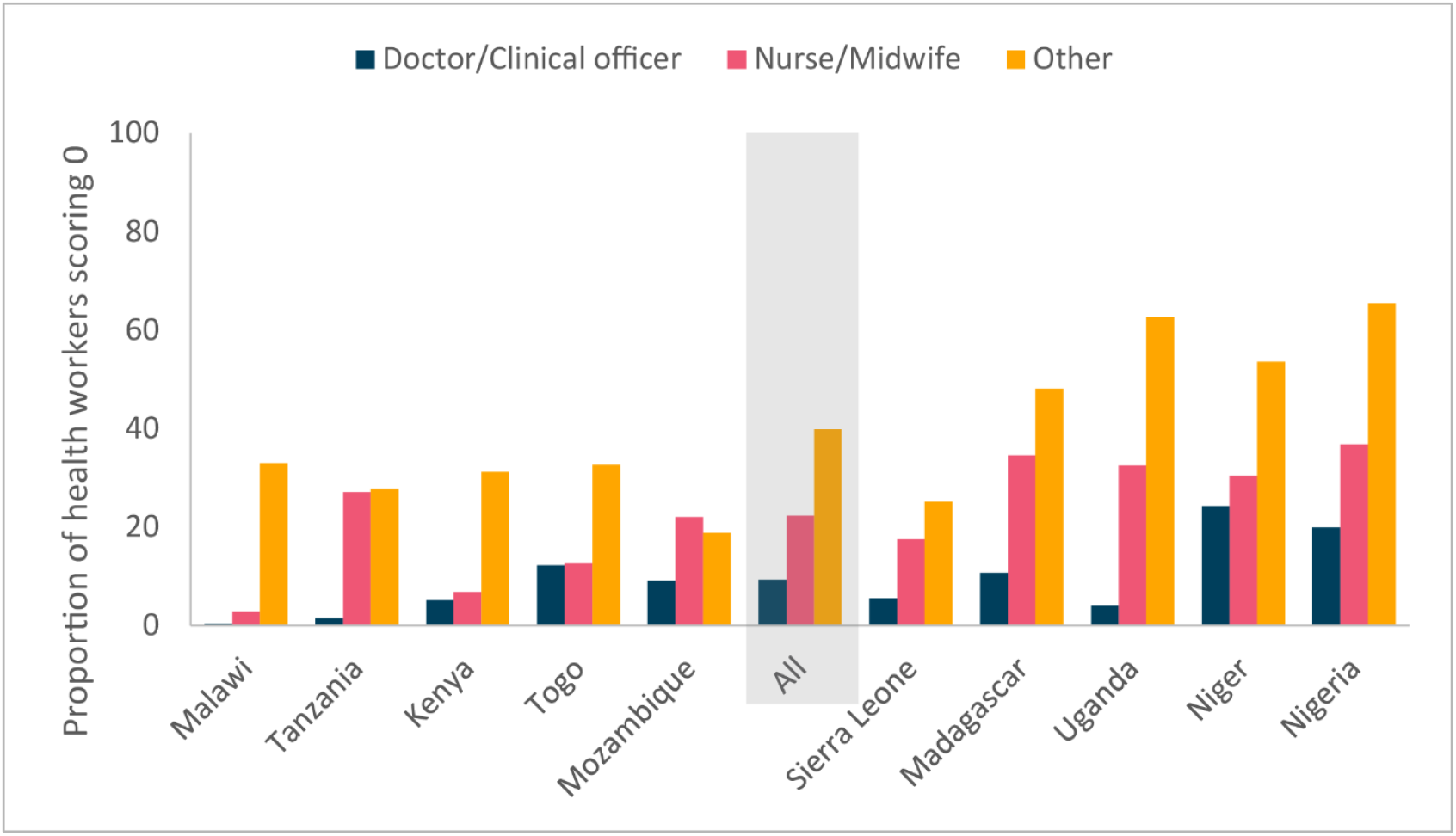
Proportion of health workers with a score of zero on a series of vignettes

Finally, we explored the relationship between diagnostic and treatment accuracy and facility and health worker characteristics. The covariates in the full model explained 42% of the variance seen in health worker diagnostic and treatment accuracy. The facility level covariates that were statistically significantly associated with diagnostic and treatment accuracy include facility type, managing authority, urbanicity, and infrastructure availability. Health worker diagnostic and treatment accuracy was lower for health workers at health centers (β = -5.193, p-value <0.001) and health posts (β = -6.977, p-value <0.001) as compared to health worker at hospitals. However, health worker diagnostic and treatment accuracy was higher for health workers at rural facilities (β = 1.488, p-value = 0.009) compared to urban facilities, for health workers at public facilities (β = 4.901, p-value <0.001) compared to private/NGO facilities, and for facilities with higher levels of infrastructure availability (β = 0.026, p-value = 0.006). Interestingly, equipment availability was the only facility level covariate not statistically significantly associated with health worker diagnostic and treatment accuracy, thus not supporting the idea that more competent health workers choose to work in better equipped facilities. The health worker level covariates that were statistically significantly associated with diagnostic and treatment accuracy include cadre, age, sex, and education. Health worker diagnostic and treatment accuracy was lower for nurses (β = -7.813, p-value <0.001) and other workers (β = -16.207, p-value <0.001) as compared to doctors as well as for health workers over the age of 50 (β = -1.8313 p-value = 0.007) as compared to those under the age of 30. In addition, health worker diagnostic and treatment accuracy was higher for male health workers (β = 4.579, p-value <0.001) as compared to female health workers and for health workers with post-secondary education (β = 5.622, p-value <0.001) as compared to those with only a primary education (Table 10). Our findings demonstrate that health workers in health centers and health posts have lower competency than those in hospitals which means the least qualified health workers are at the frontline of primary health care. In addition, the public sector draws competent health workers as evidenced by the higher competency of public sector health workers as compared to private sector health workers. Finally, rural health workers have slightly higher competency, as do men and younger health workers.

## 4. Discussion

### 4.1. Key findings

This study utilized SDI data to explore health workforce challenges across and within ten countries in Sub-Saharan Africa. We found that on average, health centers and health posts have between 2.5 and 3 clinical health workers — well below the expected staffing norms— but that there are many health centers or health posts with one or no clinical health workers. Thus, even if health facilities are reasonably staffed on average, this masks significant shortages within countries. We also found high levels of absence across all types of health facilities, driven by a range of factors. Crucially, effective staffing fell dramatically when absence was incorporated. Absence was highest at the smallest facilities and at public facilities. We observed large variation in caseloads, including several countries with per worker daily caseloads of under 16 (i.e., one or fewer cases per half hour in an eight-hour shift). Facilities in rural and urban areas had similar caseloads per effective clinical health worker. This suggests that in many settings, at current patient levels, there is no shortage in the quantity of health workers. Furthermore, the large variation within countries suggests that much of the shortage may be due to inefficiencies or challenges in deployment. While staffing availability may not be a major constraint in many settings, can patients get the care they need? We found that a significant proportion of medical workers scored zero on a series of vignettes about important maternal and child health conditions, from 0.4 percent of doctors and clinical officers in Malawi to 24.3 percent in Niger, with higher numbers for nurses and midwives in each country as compared to doctors.

Health staffing challenges in African health systems are complex, multi-faceted, and unique to individual country contexts, as demonstrated by the significant between- and within-country variation across all human resources for health performance measures. However, some challenges are ubiquitous: health workers lack basic clinical competencies, caseloads are imbalanced, and there is substantial absence of workers from health facilities; all of these contribute to sub-standard service delivery to patients and inefficient health systems. With expanding populations and large unmet need, health systems in Africa will require a significant expansion of the health workforce. However, widespread staffing shortages do not appear to be the principal constraint in the short term.

The study complements previous analyses of the SDI surveys. Di Giorgio et al (2020) focused on overall health center readiness, finding that—on average across countries—the probability that a patient visiting a health center would find at least one staff present with both the competency and essential inputs required to provide child, neonatal and maternity care is just 14 percent. Andrews et al. (2021) demonstrated the variation in the quality of care both between and within communities, which is substantial. Gatti et al. (2021) provided an overview analysis of health worker absence, caseloads, health worker ability, and infrastructure (including equipment, supplies, and medicine). This study adds a comprehensive focus on health workers, including a demonstration of where the data show dramatic staffing gaps (e.g., facilities with only one or even zero clinical health workers) and adjusting staffing and caseloads for health worker absence, which significantly shifts the results relative to previous work.

### 4.2. Implications of these findings

Three key findings from this study merit highlighting as they have substantial implications for improving health outcomes. First, primary health care facilities, the backbone of universal health coverage (UHC) and a primary access point for the most vulnerable population groups, are the most affected by health workforce challenges. Many frontline facilities lack clinical staff and have the least qualified health workers. This has important implications for the quality of primary care and the way health systems are designed. Only when high quality primary care is available can systems build mechanisms to improve the efficiency of the health sector.

Second, although there is noteworthy variation across countries and health worker cadres, the starkest finding is the low level of competence of health workers. This has implications for both the stock and flow of health workers. For existing cadres, creative solutions are needed to improve the quality of care, including new models for in-service training, use of technology to support diagnosis and treatment, and supportive supervision. These findings also suggest that there is a crisis in medical and nursing education. Addressing this will also require systemic changes, including reforms of curricula and pedagogical approaches, leveraging technology, improved oversight of both public and private training institutions, and stricter competency requirements for graduating students. A related implication of our findings is that given the low caseload, a health workforce strategy cannot be devised in isolation of understanding and addressing the reasons for low levels of utilization of health services, including poor physical access, financial barriers, low quality, and lack of trust.

Finally, we did not see a single facility type or health worker cadre have consistently higher absenteeism, and unauthorized absence was very low (under 10%). These findings indicate that high absenteeism is a human resource management issue that must be improved if countries are to provide the best possible quality care to patients. While activities beyond providing health services at the health facility are an important part of a health worker’s job, facilities must ensure that quality care can be provided to patients in need.

### 4.3. Limitations of this work

Some limitations of this study warrant discussion. First, we used health worker caseload as a proxy for workload, acknowledging that this is an imperfect measure of each health worker’s workload, especially in smaller health facilities where health workers are often in charge of administrative tasks. Second, the differences in the way health systems are organized across countries may limit cross-country comparability of this measure. Nevertheless, the large variation in caseload does highlight significant differences which cannot be solely explained by health system differences. Third, to assess health workers’ clinical competency, we use clinical vignettes, which also present some limitations: they assess what a health worker *knows* about the selected clinical *scenarios* but fail to capture what health workers *do in practice* (Das et al., 2008, Leonard and Masatu, 2010). There are trade-offs when it comes to selecting the best methodology to assess health workers’ clinical competency, as some more advanced methods—such as the use of standardized patients—are not suitable for nationally representative studies at scale that allow for cross country comparison. Hence, the use of clinical vignettes is a useful methodology to enable measurement, monitoring and comparability of health workers’ clinical competencies. Finally, our analysis consists of a series of descriptive statistics and correlations. It can serve as a complement to quasi-experimental and experimental work to understand how changing factors in health systems translate into better outcomes for patients.

## 5. Conclusions

This analysis demonstrates how SDI data may be useful to explore health workforce challenges. It also points to future research that can help countries develop evidence-informed health workforce strategies and policies; such as the need to better understand the low demand for health care and the bottlenecks to improve health workers’ clinical capacity, combined with the role played by human resource policies and regulations, and real world decision making on staffing presence/absence and how these impact the demand for care and the quality of care received by patients, among others. It also highlights the need for nimble approaches to routinely monitor health worker performance through fast and cost-effective data collection and analysis methods to provide timely information to decision makers. Such timely data is also paramount to better identify effective interventions to improve the availability and quality of health workers in the region and monitor progress over time against pre-determined health worker performance objectives. Future efforts are needed to further study staffing challenges in Sub-Saharan Africa including investigating (i) the know-do gap; (ii) the cyclical causality of low staffing, high absenteeism, low competency, and low demand/caseload levels; and (iii) the best interventions to reduce health worker absence and increase competency overall, both by improving the production and retainment of health workers.

As countries continue to strive for UHC, solutions to health staffing challenges will become increasingly vital for success. Our findings highlight the need to shift focus from simply counting the number of health workers to routinely monitoring the number of *skilled health workers working efficiently to deliver quality health services to the population* to make substantial gains towards solving the health workforce crisis in Africa. There is an opportunity for countries to reconfigure health systems for greater efficiency and quality, but this will require a greater evidence base for country planning as each country faces unique health workforce challenges.

## Data Availability

The datasets generated and/or analyzed during the current study are available in the World Bank data repository, https://microdata.worldbank.org/index.php/catalog/sdi

## Author contributions

LD, DE, ML, and AS conceptualized and designed the study. AS performed the data analysis with support from RC. All coauthors contributed to the interpretation of the findings. LD, DE, and AS wrote the initial draft of the manuscript and all co-authors contributed to revising it. All co-authors approved the final version of the manuscript and accept responsibility to submit it for publication.

## Reflexivity Statement

The authors include six females and five males and span multiple levels of seniority. The authors are based in academic institutions, an international organization, and a nonprofit think tank. The authors are a multicultural, multinational team, including an author who is Senegalese and based in Côte d’Ivoire, and all authors have extensive experience working, including conducting Service Delivery Indicator surveys, in sub-Saharan Africa. Our authors do not include an individual from each LMIC represented in this analysis as this work was conducted through the World Bank headquarters to explore cross-country findings from the Service Delivery Indicators surveys.

## Ethical approval

Each of the SDI surveys were carried out in collaboration with the Ministry of Health in each country. All survey participants gave consent to participate in the survey. Details regarding the ethics approval are available in each country survey report. The current study is a secondary analysis of Service Delivery Indicators data.

## Acknowledgements

The authors thank the many data collectors, survey experts, researchers, and country partners who have worked on the country surveys that made this cross-country paper possible. We are grateful to the World Bank, and in particular the Service Delivery Indicators Trust Fund (funded in large part by the Hewlett Foundation) for supporting the underlying data collection, of which this paper reports secondary analysis. Svensson also gratefully acknowledges financial support from Handelsbankens forskningsstiftelser. The authors thank participants at the International Health Economic Associations 2021 meeting for suggestions. The findings, interpretations, and conclusions expressed in this paper are entirely those of the authors. They do not necessarily represent the views of the World Bank and its affiliated organizations, or those of the Executive Directors of the World Bank or the governments they represent.

## Supplementary Information

**Supplementary Table S1:**
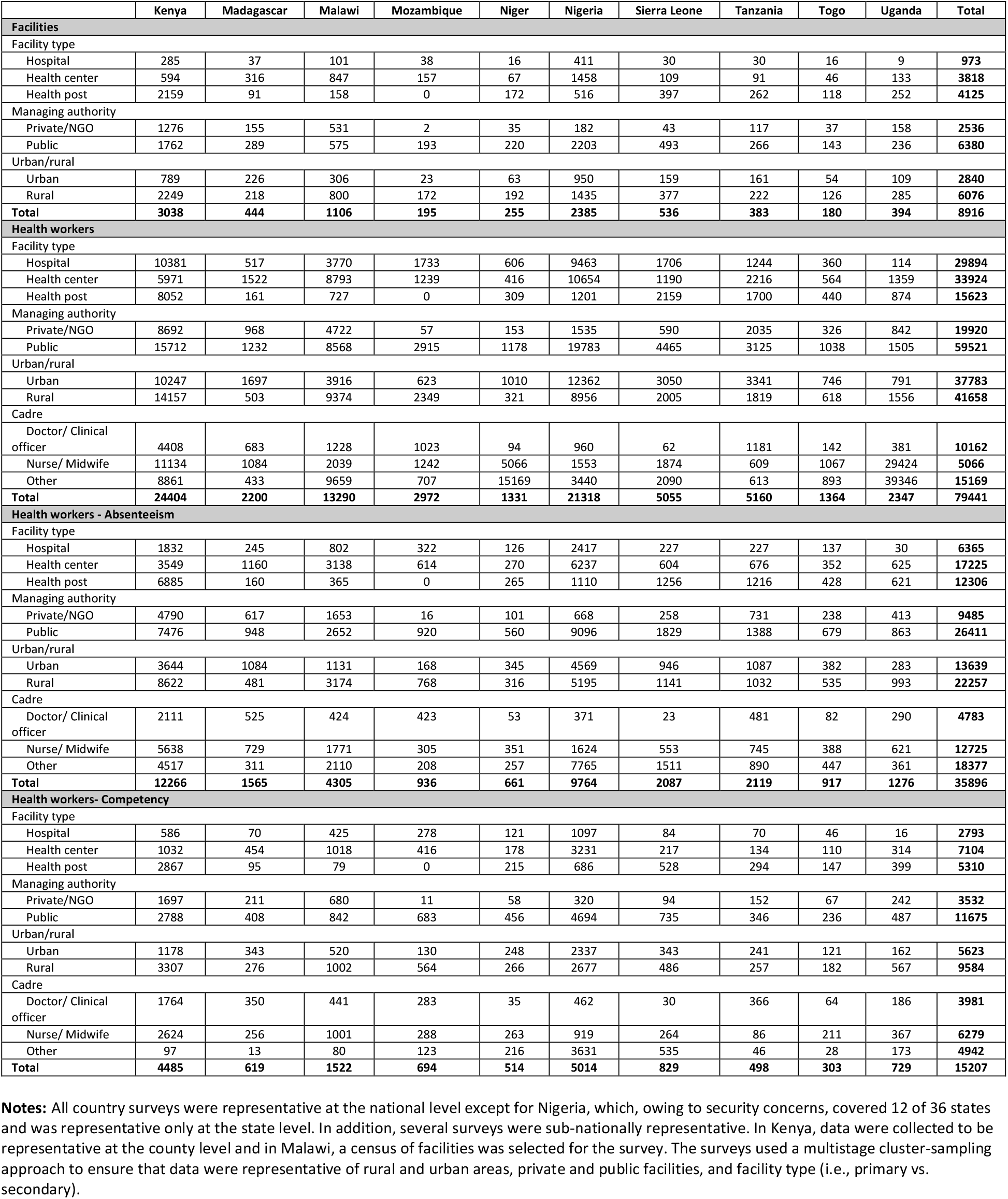
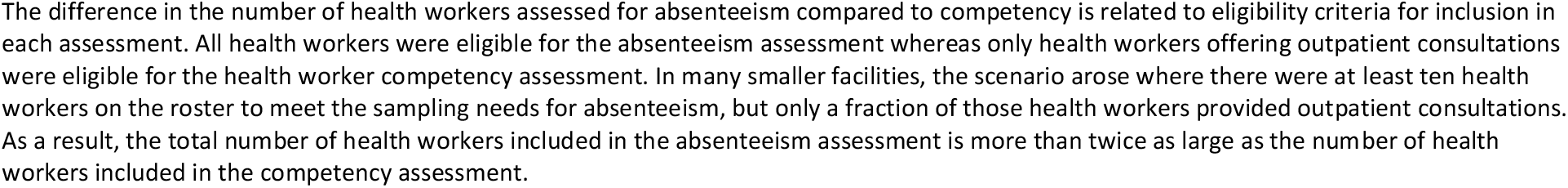
Total number of facilities, health workers, health workers assessed for absenteeism, and health workers assessed for competency, by facility type, managing authority, urban/rural, cadre, and country.

**Supplementary Table S2:** Diagnostic accuracy and treatment accuracy detailed definitions.

| Disease | Diagnostic accuracy | Treatment accuracy |
| --- | --- | --- |
| Diarrhea with dehydration | The integrated management of childhood illness (IMCI) guidelines suggest that this case should be classified as diarrhea with severe dehydration due to the presence of three warning signs: lethargy, sunken eyes, and skin pinch going back very slowly.<sup>14</sup> Clinicians should arrive at the dual diagnosis of diarrhea and dehydration but are allowed a broader range of classifications than suggested by the IMCI guidelines. These diagnoses include any mention of diarrhea (“diarrhea” or “acute diarrhea”) and any mention of dehydration (“dehydration,” “moderate dehydration,” or “severe dehydration”). In Mozambique and Niger, the child was listed as unable to drink or drinking poorly, another risk sign for severe dehydration. The way that the diagnosis for diarrhea with dehydration was recorded varied slightly from country-to country. For example, Nigeria and Uganda listed “acute diarrhea with severe dehydration” as the only possible diagnosis. Madagascar listed “diarrhea with moderate dehydration” and “diarrhea with severe dehydration.” Togo listed “diarrhea with severe dehydration.” All of these answers are counted as correct. For these countries, it is not possible to provide an accurate estimate of how many doctors diagnosed diarrhea alone; it is only possible to calculate the joint diagnosis rate. | The World Health Organization (WHO) guidelines on the integrated management of childhood illness (IMCI) note that the correct treatment of diarrhea with severe dehydration is to give intravenous fluid immediately, to insert a nasogastric (NG) tube if that is not possible, and to refer the patient to a higher-level facility if neither treatment is available. If the child has only some dehydration, then oral rehydration salts (ORS) are the recommended treatment. Given the symptoms, the correct treatment should be rehydration with an intravenous (IV) line or an NG tube. However, the use of ORS plus zinc is also counted as correct. Because the child was able to drink in most vignettes, the providers may have incorrectly believed that the dehydration was less severe. Correct treatment rates would be much lower if only IV fluids or an NG tube was counted as correct (13% correct). Uganda did not include an option for ORS, so only treatment with IV fluids or an NG tube is taken as a correct response. Kenya did not include an option for IV fluids or NG tube, so only ORS with zinc is taken as a correct response. |
| Pneumonia | A diagnosis of pneumonia is counted as the correct response. | The IMCI guidelines suggest oral amoxicillin for five days as treatment for pneumonia. Severe pneumonia can be treated with “the first dose of an appropriate antibiotic” and urgent referral to a hospital. In addition, children had a fever of 38.5 in the vignette, and |

<sup>14</sup> World Health Organization. 2014. “IMCI Chart Booklet.” Geneva, Switzerland. https://www.who.int/docs/default-source/mca-documents/imci-chart-booklet.pdf?sfvrsn=f63af425_1&download=true.

| Disease | Diagnostic accuracy | Treatment accuracy |
| --- | --- | --- |
|  |  | IMCI guidelines recommend an antipyretic in this case. Correct treatment is counted as treatment with amoxicillin, a first-line antibiotic, and any antipyretic. |
| Diabetes | Based on the characteristics presented in this vignette, a diagnosis of type 2 diabetes is the correct response. However, the option of diabetes (type not specified) was available in Kenya, Madagascar, Mozambique, Niger, Sierra Leone, and Tanzania. Clinicians were not prompted to select a specific type if they answered diabetes. Although treatment varies for different types of diabetes, the general diabetes response is also classified as correct. This practice results in higher correct diagnosis rates, an improvement by 14–50 percentage points depending on the country. | The WHO package of essential noncommunicable disease interventions (PEN) protocols states, “Individuals with persistent fasting blood glucose >6 mmol/l despite diet control should be given metformin and/or insulin as appropriate.”<sup>15</sup> Correct treatment is counted as any hypoglycemic (including insulin) or referral to a specialist. Referral to a higher level is the recommended protocol for diabetes at the primary level in multiple countries so that option is counted as correct. Although PEN protocol suggests diet control before prescribing hypoglycemics, here prescriptions on first presentation are counted as correct. |
| Tuberculosis | A diagnosis of tuberculosis is counted as the correct response. | The WHO guidelines for treatment of tuberculosis recommend combination therapy, ideally with a fixed-dose combination.<sup>16</sup> Providers are simply required to mention combination therapy. Knowledge of correct duration and dosage is not necessary, and providers would score worse if this knowledge were required. For example, 23% of providers prescribed combination therapy, but only 8% accurately recalled the correct dosage and timing (this comparison is possible in Madagascar, Mozambique, Niger, Sierra Leone, and Tanzania). However, Nigeria and Uganda recorded “correct duration and dose” as one option, so providers are assessed on having gotten the correct dosage and timing. This likely creates a downward bias for provider treatment abilities in |

<sup>15</sup> World Health Organization. 2019. “WHO Package of Essential Noncommunicable (PEN) Disease Interventions: Noncommunicable Diseases and Their Risk Factors.” Geneva, Switzerland. www.who.int/ncds/management/pen_tools/en/.
<sup>16</sup> World Health Organization. 2018. “Guidelines for Treatment of Drug-Susceptible Tuberculosis and Patient Care (2017 Update).” Geneva, Switzerland. www.who.int/tb/publications/2017/dstb_guidance_2017/en/

| Disease | Diagnostic accuracy | Treatment accuracy |
| --- | --- | --- |
|  |  | these two countries. Kenya did not record any information on whether providers got the correct duration and dosage. |
| Malaria with anemia | This case should be classified as malaria with anemia, and clinicians are required to arrive at this dual diagnosis. All countries except Togo included “malaria” as an option, and all countries except Nigeria and Uganda included “simple malaria” as an option. Both of these diagnoses are counted as correct. All countries also included “severe malaria” as an option, and this nonspecific diagnosis is not counted as correct because the case does not meet the definition of severe, and severe malaria would require different treatment. Anemia was listed simply as “anemia” without specifying severity. Providers therefore received credit for specifying malaria or simple malaria and anemia. Kenya excluded this module entirely, so it is omitted and its diagnostic accuracy is counted as the average of the four other vignettes. | IMCI guidelines recommend that children with a positive malaria test should be given “recommended first-line antimalarial” and “one dose of paracetamol in clinic” for fever reduction. In addition, iron should be given for treatment of anemia. The questions on malaria treatment varied a bit between countries, and credit is given for treatment with any artemisinin combination therapy or artemether-lumefantrine (coartem). In addition to antimalarials, the provider must prescribe paracetamol and iron for the anemia. Kenya did not include the malaria vignette and is excluded. Nigeria and Uganda did not include questions about iron and are excluded for the sake of comparability. |

**Supplementary Table S3:** Additional facility level measures detailed definitions.

| Domain | Item | Definition |
| --- | --- | --- |
| Infrastructure | Improved sanitation | Credit is given if facility reports and enumerator confirms facility has one or more functioning flush toilets or ventilated improved pit (VIP) latrines, or covered pit latrine (with slab). |
| Infrastructure | Improved water source | Credit is given if facility reports their main source of water is piped into the facility, piped onto facility grounds, or comes from a public tap/standpipe, tubewell/borehole, a protected dug well, a protected spring, bottled water, or a tanker truck. This definition is based on the WHO/UNICEF Joint Monitoring Program for Water Supply, Sanitation and Hygiene. |
| Infrastructure | Electricity | Credit is given if facility reports using electric power grid, fuel-operated generator, battery-operated generator, or a solar powered system as their main source of electricity. |
| Equipment | Thermometer | Credit is given if a facility reports and the enumerator observes that the facility has one or more functioning thermometers (used for measuring patient body temperature). |
| Equipment | Stethoscope | Credit is given if a facility reports and the enumerator observes that the facility has one or more functioning stethoscopes. |
| Equipment | Sphygmomanometer | Credit is given if a facility reports and the enumerator observes that the facility has one or more functioning sphygmomanometers. |
| Equipment | Weighing scale | Credit is given if a facility reports and the enumerator observes that the facility has one or more functioning adult, child or infant weighing scale. |

**Supplementary Table S4:** Staffing norms by cadre of clinical staff and health facility type.

|  |  | Doctor | Clinical officer | Doctor/clinical officer | Nurse/Midwife | Nurse | Midwife | Total |
| --- | --- | --- | --- | --- | --- | --- | --- | --- |
| Kenya | Health center | 2 | 11 |  | 37 |  |  | 50 |
|  | Dispensary | 0 | 2 |  | 8 |  |  | 10 |
| Malawi | Urban health center |  |  | 12 | 14 |  |  | 26 |
|  | Rural health center |  |  | 3 | 8 |  |  | 11 |
| Nigeria | Health center | 0 | 0 |  | 3 |  |  | 3 |
| Niger | Health center II | 1 |  |  | 6 |  |  | 7 |
|  | Health center I | 0 |  |  | 4 |  |  | 4 |
|  | Health post | 0 |  |  | 1 |  |  | 1 |
| Uganda | Health center IV | 2 | 4 |  | 11 |  |  | 17 |
|  | Health center III | 0 | 2 |  | 6 |  |  | 8 |
|  | Health center II | 0 | 0 |  | 2 |  |  | 2 |
| Tanzania | Health center (minimum number) | 2 | 2 |  | 10 |  |  | 14 |
|  | Health center (maximum number) | 2 | 3 |  | 15 |  |  | 20 |
|  | Dispensary (minimum number) | 0 | 1 |  | 4 |  |  | 5 |
|  | Dispensary (maximum number) | 0 | 2 |  | 7 |  |  | 9 |
| Togo | Primary healthcare center I | 0 |  |  |  | 2 | 1 | 3 |
|  | Primary healthcare center II | 1 |  |  |  | 4 | 3 | 8 |

**Notes:** Roman numerals after facility types refer to the size of the facility (i.e., health center II is larger than health center I). Staffing norms were not available for all countries.
**Sources:** These numbers are drawn from the following documents (see the reference list at the end of this supplementary appendix for more information): Kenya Ministry of Health (2014); Malawi Ministry of Health and Population (2018); Nigeria Federal Ministry of Health (2007); Ministère de la Santé Publique du Niger (2016); Uganda Ministry of Public Service (No date); Tanzania Ministry of Health and Social Welfare (2013); Ministère de la Santé du Togo (2013).

**Supplementary Table S5:** Average staffing norms compared to actual staffing levels.

|  | EXPECTED average<br>number of clinical<br>staff | ACTUAL average<br>number of clinical<br>staff in HCs and HPs |
| --- | --- | --- |
| Kenya | 30 | 3.35 |
| Malawi | 18.5 | 3.37 |
| Nigeria | 3 | 0.58 |
| Niger | 5.5 | 1.38 |
| Uganda | 12.5 | 3.42 |
| Togo | 5.5 | 2.6 |
| Tanzania | 12 | 4.36 |

**Notes:** The “expected” average number of clinical staff is the average across cadres and facility types from Supplementary Table S4. The “actual” number of clinical staff is from Table 3.
**Source:** Authors’ construction.

**Supplementary Table S6:** Health worker absenteeism - total, by country (% and 95% CI)

|  | Kenya | Madagascar | Malawi | Mozambique | Niger | Nigeria | Sierra Leone | Tanzania | Togo | Uganda | All |
| --- | --- | --- | --- | --- | --- | --- | --- | --- | --- | --- | --- |
| National | 53.8<br>(52.0 - 55.6) | 26.0<br>(22.5 - 29.6) | 36.5<br>(34.0 - 39.1) | 23.3<br>(19.0 - 27.6) | 31.1<br>(26.4 - 35.9) | 34.4<br>(32.5 - 36.3) | 39.9<br>(36.8 - 42.9) | 13.8<br>(11.0 - 16.5) | 43.8<br>(37.1 - 50.5) | 43.8<br>(38.7 - 48.9) | 34.7<br>(26.3 - 43) |
| Facility type |  |  |  |  |  |  |  |  |  |  |  |
| Hospital | 58.9<br>(55.0 - 62.8) | 19.2<br>(13.0 - 25.5) | 43.3<br>(37.6 - 49.0) | 23.9<br>(18.0 - 29.8) | 38.2<br>(27.1 - 49.4) | 30.3<br>(26.6 - 34.1) | 34.0<br>(26.4 - 41.5) | 12.6<br>(6.6 - 18.6) | 47.6<br>(38.9 - 56.4) | 27.7<br>(6.0 - 49.4) | 33.6<br>(23.6 - 43.5) |
| Health center | 52.3<br>(50.0 - 54.6) | 29.1<br>(24.3 - 33.9) | 34.3<br>(32.2 - 36.5) | 22.3<br>(16.7 - 27.9) | 35.0<br>(28.1 - 42.0) | 38.2<br>(36.5 - 39.9) | 39.0<br>(33.2 - 44.8) | 14.9<br>(9.4 - 20.3) | 40.9<br>(31.1 - 50.8) | 46.2<br>(38.9 - 53.5) | 35.2<br>(27.4 - 43.1) |
| Health post | 48.5<br>(47.0 - 49.9) | 24.0<br>(14.3 - 33.6) | 9.2<br>(5.4 - 12.9) | NA | 20.4<br>(13.5 - 27.3) | 32.5<br>(29.3 - 35.6) | 43.0<br>(39.1 - 46.9) | 13.7<br>(11.2 - 16.3) | 45.9<br>(35.7 - 56.1) | 43.5<br>(37.6 - 49.4) | 31.2<br>(19.8 - 42.6) |
| Managing authority |  |  |  |  |  |  |  |  |  |  |  |
| Private/NGO | 46.3<br>(43.6 - 49.0) | 25.5<br>(20.4 - 30.5) | 30.7<br>(27.8 - 33.7) | 11.4<br>(0.0 - 30.2) | 34.7<br>(22.2 - 47.2) | 19.6<br>(15.4 - 23.7) | 33.9<br>(26.6 - 41.3) | 11.9<br>(8.0 - 15.8) | 44.5<br>(33.2 - 55.8) | 38.7<br>(31.0 - 46.5) | 29.7<br>(20.8 - 38.6) |
| Public | 58.6<br>(56.2 - 60.9) | 26.3<br>(21.6 - 31.1) | 39.2<br>(35.8 - 42.7) | 23.6<br>(19.2 - 27.9) | 30.9<br>(25.9 - 35.9) | 38.9<br>(37.0 - 40.8) | 40.8<br>(37.5 - 44.1) | 14.9<br>(11.2 - 18.6) | 43.0<br>(35.8 - 50.3) | 47.5<br>(40.9 - 54.2) | 36.4<br>(27.3 - 45.5) |
| Urban/Rural |  |  |  |  |  |  |  |  |  |  |  |
| Urban | 55.1<br>(51.8 - 58.4) | 24.5<br>(21.0 - 28.1) | 43.1<br>(37.5 - 48.7) | 23.2<br>(13.2 - 33.2) | 34.8<br>(28.0 - 41.5) | 33.4<br>(30.5 - 36.3) | 37.8<br>(33.5 - 42.0) | 14.3<br>(10.3 - 18.2) | 45.5<br>(35.9 - 55.1) | 36.5<br>(26.9 - 46.1) | 34.8<br>(26.3 - 43.3) |
| Rural | 52.6<br>(50.8 - 54.4) | 27.7<br>(21.3 - 34.2) | 32.5<br>(30.4 - 34.6) | 23.4<br>(19.0 - 27.9) | 27.6<br>(21.1 - 34.2) | 35.8<br>(33.8 - 37.8) | 42.7<br>(38.5 - 46.9) | 12.8<br>(10.1 - 15.4) | 41.0<br>(32.7 - 49.3) | 49.3<br>(45.0 - 53.7) | 34.6<br>(25.8 - 43.3) |
| Health worker cadre |  |  |  |  |  |  |  |  |  |  |  |
| Doctor/<br>Clinical officer | 56.1<br>(51.8 - 60.5) | 32.6<br>(25.5 - 39.6) | 41.4<br>(31.8 - 51.0) | 24.3<br>(18.2 - 30.4) | 38.3<br>(20.4 - 56.2) | 28.9<br>(21.4 - 36.5) | 24.9<br>(3.9 - 46.0) | 21.1<br>(13.6 - 28.6) | 58.2<br>(37.0 - 79.4) | 42.5<br>(31.7 - 53.3) | 36.8<br>(27.6 - 46.1) |
| Nurse /<br>Midwife | 55.8<br>(52.9 - 58.8) | 24.9<br>(19.9 - 29.9) | 33.1<br>(29.2 - 37.0) | 24.7<br>(17.3 - 32.1) | 32.3<br>(25.9 - 38.6) | 38.7<br>(33.1 - 44.4) | 31.9<br>(26.2 - 37.6) | 10.5<br>(7.4 - 13.5) | 48.4<br>(38.3 - 58.5) | 47.7<br>(41.9 - 53.5) | 34.8<br>(25.2 - 44.4) |
| Other worker | 49.6<br>(47.2 - 52.1) | 18.5<br>(12.8 - 24.1) | 38.8<br>(36.2 - 41.3) | 18.8<br>(11.5 - 26.2) | 28.2<br>(20.5 - 35.9) | 33.6<br>(31.9 - 35.3) | 43.7<br>(40.0 - 47.3) | 12.3<br>(7.9 - 16.6) | 36.5<br>(26.9 - 46.0) | 38.3<br>(32.1 - 44.5) | 31.8<br>(23.2 - 40.5) |

**Supplementary Table S7:**
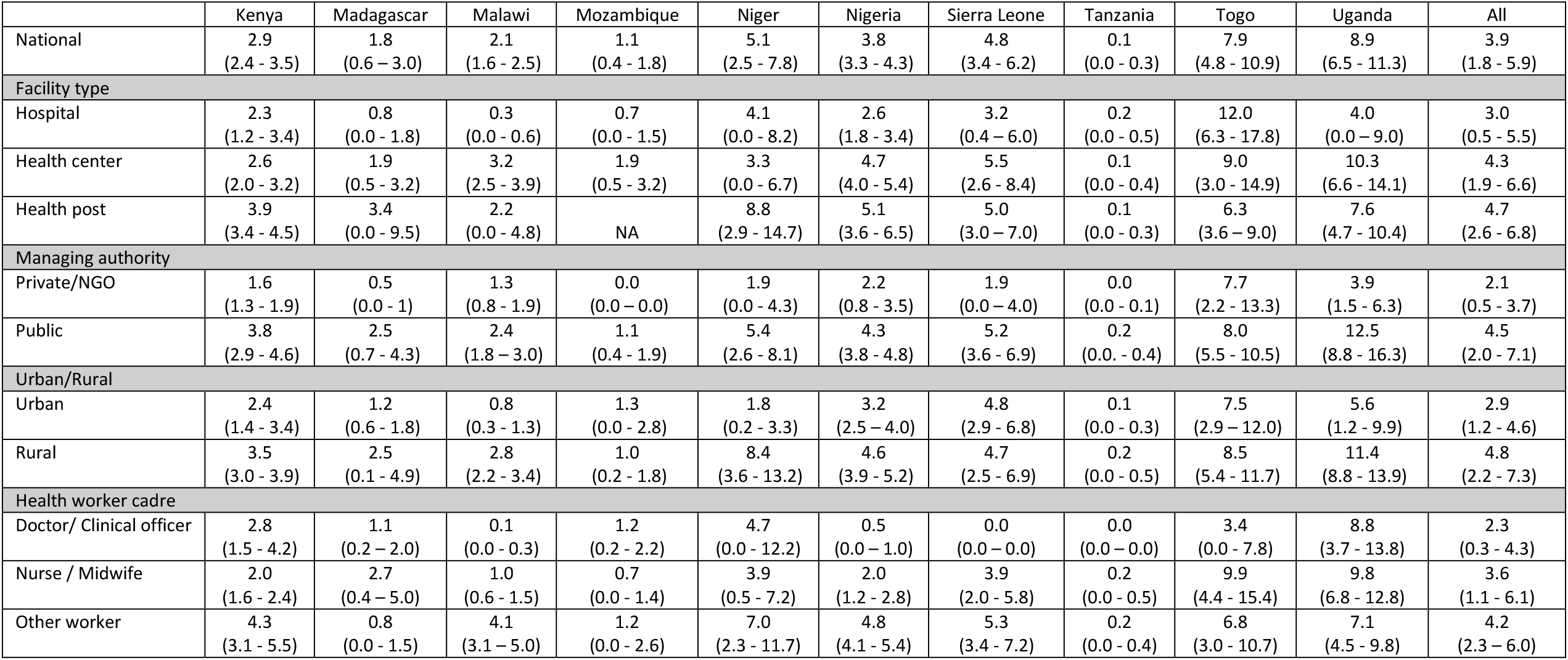
Health worker absenteeism - unauthorized, by country (% and 95% CI)

|  | Kenya | Madagascar | Malawi | Mozambique | Niger | Nigeria | Sierra Leone | Tanzania | Togo | Uganda | All |
| --- | --- | --- | --- | --- | --- | --- | --- | --- | --- | --- | --- |
| National | 2.9<br>(2.4 - 3.5) | 1.8<br>(0.6 - 3.0) | 2.1<br>(1.6 - 2.5) | 1.1<br>(0.4 - 1.8) | 5.1<br>(2.5 - 7.8) | 3.8<br>(3.3 - 4.3) | 4.8<br>(3.4 - 6.2) | 0.1<br>(0.0 - 0.3) | 7.9<br>(4.8 - 10.9) | 8.9<br>(6.5 - 11.3) | 3.9<br>(1.8 - 5.9) |
| Facility type |  |  |  |  |  |  |  |  |  |  |  |
| Hospital | 2.3<br>(1.2 - 3.4) | 0.8<br>(0.0 - 1.8) | 0.3<br>(0.0 - 0.6) | 0.7<br>(0.0 - 1.5) | 4.1<br>(0.0 - 8.2) | 2.6<br>(1.8 - 3.4) | 3.2<br>(0.4 - 6.0) | 0.2<br>(0.0 - 0.5) | 12.0<br>(6.3 - 17.8) | 4.0<br>(0.0 - 9.0) | 3.0<br>(0.5 - 5.5) |
| Health center | 2.6<br>(2.0 - 3.2) | 1.9<br>(0.5 - 3.2) | 3.2<br>(2.5 - 3.9) | 1.9<br>(0.5 - 3.2) | 3.3<br>(0.0 - 6.7) | 4.7<br>(4.0 - 5.4) | 5.5<br>(2.6 - 8.4) | 0.1<br>(0.0 - 0.4) | 9.0<br>(3.0 - 14.9) | 10.3<br>(6.6 - 14.1) | 4.3<br>(1.9 - 6.6) |
| Health post | 3.9<br>(3.4 - 4.5) | 3.4<br>(0.0 - 9.5) | 2.2<br>(0.0 - 4.8) | NA | 8.8<br>(2.9 - 14.7) | 5.1<br>(3.6 - 6.5) | 5.0<br>(3.0 - 7.0) | 0.1<br>(0.0 - 0.3) | 6.3<br>(3.6 - 9.0) | 7.6<br>(4.7 - 10.4) | 4.7<br>(2.6 - 6.8) |
| Managing authority |  |  |  |  |  |  |  |  |  |  |  |
| Private/NGO | 1.6<br>(1.3 - 1.9) | 0.5<br>(0.0 - 1) | 1.3<br>(0.8 - 1.9) | 0.0<br>(0.0 - 0.0) | 1.9<br>(0.0 - 4.3) | 2.2<br>(0.8 - 3.5) | 1.9<br>(0.0 - 4.0) | 0.0<br>(0.0 - 0.1) | 7.7<br>(2.2 - 13.3) | 3.9<br>(1.5 - 6.3) | 2.1<br>(0.5 - 3.7) |
| Public | 3.8<br>(2.9 - 4.6) | 2.5<br>(0.7 - 4.3) | 2.4<br>(1.8 - 3.0) | 1.1<br>(0.4 - 1.9) | 5.4<br>(2.6 - 8.1) | 4.3<br>(3.8 - 4.8) | 5.2<br>(3.6 - 6.9) | 0.2<br>(0.0 - 0.4) | 8.0<br>(5.5 - 10.5) | 12.5<br>(8.8 - 16.3) | 4.5<br>(2.0 - 7.1) |
| Urban/Rural |  |  |  |  |  |  |  |  |  |  |  |
| Urban | 2.4<br>(1.4 - 3.4) | 1.2<br>(0.6 - 1.8) | 0.8<br>(0.3 - 1.3) | 1.3<br>(0.0 - 2.8) | 1.8<br>(0.2 - 3.3) | 3.2<br>(2.5 - 4.0) | 4.8<br>(2.9 - 6.8) | 0.1<br>(0.0 - 0.3) | 7.5<br>(2.9 - 12.0) | 5.6<br>(1.2 - 9.9) | 2.9<br>(1.2 - 4.6) |
| Rural | 3.5<br>(3.0 - 3.9) | 2.5<br>(0.1 - 4.9) | 2.8<br>(2.2 - 3.4) | 1.0<br>(0.2 - 1.8) | 8.4<br>(3.6 - 13.2) | 4.6<br>(3.9 - 5.2) | 4.7<br>(2.5 - 6.9) | 0.2<br>(0.0 - 0.5) | 8.5<br>(5.4 - 11.7) | 11.4<br>(8.8 - 13.9) | 4.8<br>(2.2 - 7.3) |
| Health worker cadre |  |  |  |  |  |  |  |  |  |  |  |
| Doctor/ Clinical officer | 2.8<br>(1.5 - 4.2) | 1.1<br>(0.2 - 2.0) | 0.1<br>(0.0 - 0.3) | 1.2<br>(0.2 - 2.2) | 4.7<br>(0.0 - 12.2) | 0.5<br>(0.0 - 1.0) | 0.0<br>(0.0 - 0.0) | 0.0<br>(0.0 - 0.0) | 3.4<br>(0.0 - 7.8) | 8.8<br>(3.7 - 13.8) | 2.3<br>(0.3 - 4.3) |
| Nurse / Midwife | 2.0<br>(1.6 - 2.4) | 2.7<br>(0.4 - 5.0) | 1.0<br>(0.6 - 1.5) | 0.7<br>(0.0 - 1.4) | 3.9<br>(0.5 - 7.2) | 2.0<br>(1.2 - 2.8) | 3.9<br>(2.0 - 5.8) | 0.2<br>(0.0 - 0.5) | 9.9<br>(4.4 - 15.4) | 9.8<br>(6.8 - 12.8) | 3.6<br>(1.1 - 6.1) |
| Other worker | 4.3<br>(3.1 - 5.5) | 0.8<br>(0.0 - 1.5) | 4.1<br>(3.1 - 5.0) | 1.2<br>(0.0 - 2.6) | 7.0<br>(2.3 - 11.7) | 4.8<br>(4.1 - 5.4) | 5.3<br>(3.4 - 7.2) | 0.2<br>(0.0 - 0.4) | 6.8<br>(3.0 - 10.7) | 7.1<br>(4.5 - 9.8) | 4.2<br>(2.3 - 6.0) |

**Supplementary Table S8:** Reasons for health worker absenteeism, by country (% and 95% CI)

|  | Kenya | Madagascar | Malawi | Mozambique | Niger | Nigeria | Sierra Leone | Tanzania | Togo | Uganda | All |
| --- | --- | --- | --- | --- | --- | --- | --- | --- | --- | --- | --- |
| Authorized absence | 40.5<br>(37.8 - 43.2) | 35.7<br>(28.4 - 43.1) | 22.9<br>(18.7 - 27.1) | 26.3<br>(17.0 - 35.7) | 34.0<br>(25.7 - 42.4) | 32.7<br>(29.8 - 35.5) | 17.5<br>(13.8 - 21.2) | 29.6<br>(21.1 - 38.0) | 26.8<br>(18.3 - 35.3) | 24.2<br>(18.6 - 29.8) | 30.0<br>(22.5 - 37.4) |
| Official mission | 8.7<br>(7.6 - 9.8) | 13.1<br>(7.9 - 18.3) | 7.9<br>(5.1 - 10.7) | 20.8<br>(12.7 - 29.0) | 6.1<br>(2.5 - 9.7) | 15.9<br>(13.8 - 17.9) | 8.2<br>(5.7 - 10.8) | 4.0<br>(2.0 - 5.9) | 11.0<br>(3.6 - 18.5) | 11.4<br>(6.6 - 16.2) | 11.5<br>(6.6 - 16.4) |
| Outreach or fieldwork | 1.4<br>(1.1 - 1.7) | 0.2<br>(0.0 - 0.7) | 21.9<br>(19.0 - 24.8) | 0.0<br>(0.0 - 0.0) | 0.0<br>(0.0 - 0.0) | 2.8<br>(2.1 - 3.5) | 11.2<br>(9.0 - 13.4) | 50.0<br>(0.0 - 12.5) | 0.0<br>(0.0 - 0.0) | 3.5<br>(2.0 - 5.0) | 5.4<br>(2.3 - 13.1) |
| Sick or on maternity leave | 5.2<br>(4.1 - 6.3) | 5.1<br>(2.9 - 7.2) | 3.9<br>(2.4 - 5.3) | 14.7<br>(7.6 - 21.9) | 19.7<br>(12.2 - 27.1) | 11.1<br>(9.3 - 12.9) | 9.1<br>(6.9 - 11.4) | 23.5<br>(15.3 - 31.6) | 9.4<br>(2.7 - 16.0) | 7.9<br>(5.2 - 10.6) | 9.8<br>(4.4 - 15.2) |
| Training, seminar, meeting | 7.7<br>(6.2 - 9.2) | 24.8<br>(18.7 - 30.8) | 18.5<br>(14.5 - 22.5) | 21.4<br>(11.7 - 31.1) | 11.3<br>(6.0 - 16.6) | 8.8<br>(7.3 - 10.3) | 6.3<br>(3.7 - 9.0) | 35.7<br>(24.5 - 46.9) | 34.7<br>(23.6 - 45.8) | 30.6<br>(25.2 - 36.0) | 14.1<br>(7.3 - 20.9) |
| Unauthorized absence | 5.4<br>(4.5 - 6.4) | 7.1<br>(2.6 - 11.5) | 5.6<br>(4.4 - 6.9) | 4.7<br>(1.7 - 7.8) | 16.5<br>(8.8 - 24.3) | 11.4<br>(9.9 - 12.9) | 12.0<br>(8.6 - 15.4) | 1.1<br>(0.0 - 2.1) | 18.0<br>(11.1 - 24.9) | 21.9<br>(16.7 - 27.2) | 9.0<br>(4.9 - 13.1) |
| Other | 31.0<br>(28.7 - 33.3) | 14.0<br>(10.0 - 18.1) | 19.3<br>(15.2 - 23.5) | 12.0<br>(6.2 - 17.8) | 12.3<br>(6.5 - 18.2) | 17.4<br>(13.2 - 21.7) | 35.5<br>(31.5 - 39.6) | 1.2<br>(0.3 - 2.2) | 0.2<br>(0.0 - 0.4) | 0.4<br>(0.1 - 0.8) | 20.2<br>(11.6 - 28.9) |

**Supplementary Table S9:** Clinical staffing patterns in health centers and health posts adjusted for absenteeism, by country (number/proportion and 95% CI)

|  |  | Kenya | Madagascar | Malawi | Mozambique | Niger | Nigeria | Sierra Leone | Tanzania | Togo | Uganda | All |
| --- | --- | --- | --- | --- | --- | --- | --- | --- | --- | --- | --- | --- |
|  |  | n = 2753 | n = 407 | n = 1005 | n = 157 | n = 239 | n = 1974 | n = 506 | n = 353 | n = 164 | n = 385 | n = 7943 |
| Proportion of health centers and health posts with one or fewer clinical staff adjusted for absenteeism | A: Total number of clinical staff reduced by proportion of clinical staff absent | 63.9<br>(61.9 - 65.8) | 69.2<br>(61.4 - 77.0) | 39.4<br>(36.4 - 42.4) | 47.8<br>(39.9 - 55.6) | 85.3<br>(80.4 - 90.3) | 92.4<br>(91.0 - 93.7) | 84.4<br>(80.7 - 88.2) | 31.0<br>(24.3 - 37.8) | 68.9<br>(54.3 - 83.5) | 59.0<br>(52.8 - 65.2) | 64.1<br>(49.6 - 78.7) |
|  | B: Total number of clinical staff reduced by subtracting number of clinical staff absent | 63.1<br>(61.1 - 65.0) | 69.2<br>(61.5 - 77.0) | 36.2<br>(33.2 - 39.2) | 47.8<br>(39.9 - 55.6) | 85.3<br>(80.4 - 90.3) | 91.8<br>(90.4 - 93.2) | 84.2<br>(80.4 - 87.9) | 30.9<br>(24.1 - 37.7) | 68.3<br>(53.7 - 83.0) | 56.2<br>(49.9 - 62.5) | 63.3<br>(48.5 - 78.1) |
|  | B1: Subset of facilities where all clinical staff were assessed for absenteeism | n = 2238 | n = 341 | n = 117 | n = 331 | n = 97 | n = 308 | n = 206 | n = 135 | n = 183 | n = 214 | n = 4170 |
|  |  | 75.8<br>(73.9 - 77.8) | 73.2<br>(65.2 - 81.2) | 61.3<br>(56.1 - 66.6) | 56.4<br>(47.4 - 65.4) | 79.7<br>(70.3 - 89.2) | 89.8<br>(85.7 - 93.9) | 81.6<br>(75.6 - 87.6) | 41.8<br>(32.3 - 51.3) | 70.7<br>(54.4 - 87.0) | 80.0<br>(73.8 - 86.3) | 71.1<br>(60.9 - 81.2) |
| Proportion of health centers and health posts with two or fewer clinical staff adjusted for absenteeism | A: Total number of clinical staff reduced by proportion of clinical staff absent | 82.7<br>(81.2 - 84.3) | 89.3<br>(85.1 - 93.5) | 61.6<br>(58.6 - 64.6) | 67.5<br>(60.2 - 74.9) | 91.9<br>(88.4 - 95.4) | 95.6<br>(94.5 - 96.7) | 91.7<br>(88.7 - 94.8) | 60.6<br>(53.0 - 68.2) | 79.7<br>(67.3 - 92.1) | 72.7<br>(66.8 - 78.6) | 79.3<br>(70.0 - 88.7) |
|  | B: Total number of clinical staff reduced by subtracting number of clinical staff absent | 81.0<br>(79.4 - 82.6) | 88.9<br>(84.7 - 93.2) | 58.1<br>(55.1 - 61.2) | 67.5<br>(60.2 - 74.9) | 91.2<br>(87.4 - 94.9) | 94.7<br>(93.5 - 95.9) | 91.2<br>(88.1 - 94.2) | 59.8<br>(52.2 - 67.4) | 79.7<br>(67.3 - 92.1) | 68.1<br>(62.0 - 74.1) | 78.0<br>(68.2 - 87.8) |
|  | B1: Subset of facilities where all clinical staff were assessed for absenteeism | n = 2238 | n = 341 | n = 117 | n = 331 | n = 97 | n = 308 | n = 206 | n = 135 | n = 183 | n = 214 | n = 4170 |
|  |  | 93.7<br>(92.6 - 94.8) | 93.8<br>(89.9 - 97.6) | 88.5<br>(85.1 - 92.0) | 82.1<br>(75.1 - 89.0) | 92.9<br>(87.3 - 98.5) | 96.8<br>(94.0 - 99.6) | 95.7<br>(92.7 - 98.7) | 82.9<br>(76.5 - 89.3) | 83.0<br>(69.4 - 96.6) | 93.4<br>(89.8 - 97.1) | 90.3<br>(86.2 - 94.3) |

**Note:** This table presents clinical staffing adjusted for absenteeism using two alternative methods of adjustment, as outlined in endnote 5 of the main paper.

**Supplementary Table S10:** Average health worker caseload per facility, by country (number and 95% CI)

|  | Kenya | Madagascar | Malawi | Mozambique | Niger | Nigeria | Sierra Leone | Tanzania | Togo | Uganda | All |
| --- | --- | --- | --- | --- | --- | --- | --- | --- | --- | --- | --- |
| National | 23.1<br>(21.9 - 24.2) | 6.4<br>(5.0 - 7.7) | 47.5<br>(43.7 - 51.3) | 23.2<br>(20.0 - 26.5) | 10.9<br>(8.6 - 13.2) | 2.8<br>(2.5 - 3.2) | 11.3<br>(10.2 - 12.5) | 11.7<br>(9.6 - 13.8) | 6.3<br>(4.8 - 7.9) | 17.0<br>(14.2 - 19.7) | 16.0<br>(6.7 - 25.3) |
| **Facility type** |  |  |  |  |  |  |  |  |  |  |  |
| Hospital | 24.6<br>(19.9 - 29.2) | 5.5<br>(3.1 - 7.8) | 27.2<br>(19.9 - 34.5) | 16.7<br>(7.9 - 25.5) | 1.1<br>(0.6 - 1.7) | 4.1<br>(2.9 - 5.4) | 8.0<br>(6.3 - 9.8) | 33.5<br>(5.5 - 61.4) | 6.6<br>(4.0 - 9.3) | 16.6<br>(2.9 - 30.2) | 14.4<br>(6.5 - 22.3) |
| Health center | 25.0<br>(22.4 - 27.5) | 7.7<br>(5.5 - 9.9) | 49.5<br>(45.2 - 53.9) | 24.9<br>(21.5 - 28.3) | 21.6<br>(17.1 - 26.2) | 2.7<br>(2.3 - 3.0) | 11.3<br>(8.7 - 13.9) | 14.2<br>(7.6 - 20.9) | 7.6<br>(4.5 - 10.6) | 17.1<br>(11.8 - 22.4) | 18.2<br>(8.6 - 27.7) |
| Health post | 22.2<br>(21.0 - 23.5) | 4.0<br>(2.8 - 5.2) | 64.2<br>(49.1 - 79.3) | NA | 6.1<br>(4.5 - 7.6) | 2.0<br>(1.7 - 2.3) | 11.5<br>(10.1 - 12.9) | 10.4<br>(8.5 - 12.4) | 5.8<br>(4.1 - 7.6) | 16.9<br>(13.7 - 20.2) | 15.9<br>(1.1 - 30.7) |
| **Managing authority** |  |  |  |  |  |  |  |  |  |  |  |
| Private/NGO | 15.0<br>(13.5 - 16.6) | 11.8<br>(9.8 - 13.7) | 23.8<br>(19.5 - 28.1) | 12.5<br>(0.0 - 29) | 50<br>(2.6 - 7.4) | 3.2<br>(1.9 - 4.6) | 9.2<br>(7.1 - 11.3) | 11.3<br>(7.0 - 15.6) | 6.8<br>(4.3 - 9.4) | 7.0<br>(4.7 - 9.4) | 10.6<br>(6.3 - 14.8) |
| Public | 30.4<br>(28.9 - 32) | 5.1<br>(3.6 - 6.6) | 58.0<br>(53.2 - 62.8) | 23.4<br>(20.0 - 26.7) | 11.2<br>(8.7 - 13.6) | 2.7<br>(2.5 - 3.0) | 11.5<br>(10.2 - 12.8) | 11.9<br>(9.4 - 14.3) | 6.0<br>(4.0 - 8.0) | 26.1<br>(22.0 - 30.3) | 18.6<br>(6.7 - 30.6) |
| **Urban/Rural** |  |  |  |  |  |  |  |  |  |  |  |
| Urban | 20.1<br>(17.5 - 22.6) | 10.5<br>(8.7 - 12.3) | 36.7<br>(27.2 - 46.1) | 19.6<br>(5.6 - 33.6) | 8.1<br>(4.5 - 11.7) | 3.2<br>(2.5 - 3.9) | 10.5<br>(8.5 - 12.5) | 15.3<br>(9.7 - 20.9) | 6.6<br>(4.3 - 9.0) | 10.1<br>(5.7 - 14.6) | 14.1<br>(7.2 - 20.9) |
| Rural | 24.6<br>(23.4 - 25.7) | 4.9<br>(3.3 - 6.6) | 49.3<br>(45.2 - 53.5) | 23.7<br>(20.5 - 26.9) | 11.2<br>(8.7 - 13.8) | 2.6<br>(2.3 - 2.9) | 11.7<br>(10.2 - 13.2) | 10.1<br>(8.4 - 11.8) | 6.1<br>(4.0 - 8.2) | 20.2<br>(16.8 - 23.5) | 16.4<br>(6.5 - 26.4) |

**Supplementary Table S11:** Proportion of facilities with low and high caseload, by country (% and 95% CI)

|  | Kenya | Madagascar | Malawi | Mozambique | Niger | Nigeria | Sierra Leone | Tanzania | Togo | Uganda | All |
| --- | --- | --- | --- | --- | --- | --- | --- | --- | --- | --- | --- |
| **Proportion of facilities with a caseload of <5** |  |  |  |  |  |  |  |  |  |  |  |
| Private/NGO | 31.7<br>(28.7 - 34.6) | 30.7<br>(19.7 - 41.8) | 22.1<br>(15.3 - 28.8) | 50.0<br>(0 - 119.9) | 70.5<br>(52.6 - 88.3) | 83.1<br>(76.2 - 89.9) | 32.5<br>(16.8 - 48.1) | 40.5<br>(24.7 - 56.4) | 53.2<br>(23.3 - 83.2) | 64.1<br>(54.1 - 74.2) | 47.8<br>(33.6 - 62.1) |
| Public | 5.0<br>(3.8 - 6.1) | 73.1<br>(64.8 - 81.5) | 2.4<br>(0.8 - 4.1) | 13.6<br>(8.6 - 18.6) | 40.7<br>(30.3 - 51.1) | 86.5<br>(84.7 - 88.3) | 32.3<br>(27.6 - 37.1) | 35.6<br>(26.7 - 44.5) | 63.9<br>(50.2 - 77.7) | 3.4<br>(0.5 - 6.3) | 35.7<br>(13.8 - 57.5) |
| Total | 17.8<br>(16.1 - 19.4) | 65.2<br>(57 - 73.4) | 8.5<br>(5.9 - 11) | 14.0<br>(9 - 19) | 41.8<br>(31.8 - 51.8) | 85.9<br>(84 - 87.8) | 32.4<br>(27.8 - 36.9) | 37.0<br>(29.2 - 44.7) | 59.5<br>(44.9 - 74.1) | 32.5<br>(25.6 - 39.4) | 39.4<br>(21.9 - 57) |
| **Proportion of facilities with a caseload of >=30** |  |  |  |  |  |  |  |  |  |  |  |
| Private/NGO | 12.8<br>(10.5 - 15.1) | 9.1<br>(4.3 - 13.9) | 26.2<br>(19.0 - 33.4) | 0.0<br>(0.0 - 0.0) | 0.0<br>(0.0 - 0.0) | 1.9<br>(0.0 - 4.4) | 0.0<br>(0.0 - 0.0) | 3.7<br>(0.1 - 7.3) | 0.0<br>(0.0 - 0.0) | 4.7<br>(0.7 - 8.7) | 5.8<br>(0.0 - 11.8) |
| Public | 36.1<br>(33.4 - 38.7) | 1.2<br>(0.0 - 2.3) | 68.6<br>(63.6 - 73.6) | 27.2<br>(20.7 - 33.7) | 10.7<br>(4.1 - 17.2) | 0.8<br>(0.3 - 1.3) | 6.2<br>(4.1 - 8.4) | 6.5<br>(3.8 - 9.2) | 3.8<br>(0.0 - 11.2) | 29.8<br>(22.4 - 37.2) | 19.1<br>(3.6 - 34.5) |
| Total | 24.9<br>(23.1 - 26.8) | 2.6<br>(1.3 - 4.0) | 55.6<br>(51.1 - 60.1) | 26.9<br>(20.5 - 33.3) | 10.3<br>(4.0 - 16.5) | 1.0<br>(0.4 - 1.6) | 5.7<br>(3.7 - 7.7) | 5.7<br>(3.5 - 7.9) | 2.2<br>(0.0 - 6.6) | 17.8<br>(13.2 - 22.3) | 15.3<br>(3.1 - 27.4) |

**Supplementary Table S12:** Linear regression of caseload on facility characteristics.

|  | **Model 1 –**<br>**facility**<br>**characteristics on**<br>**caseload** | **Model 2 –**<br>**Model 1 +**<br>**readiness** | **Model 3 –**<br>**Model 1 +**<br>**readiness +**<br>**absenteeism** | **Model 4 –**<br>**Model 1 +**<br>**readiness +**<br>**absenteeism**<br>**+**<br>**competency** |
| --- | --- | --- | --- | --- |
| **Country (REF = Kenya)** |  |  |  |  |
| Madagascar | -15.4 *** | -14.4 *** | -10.3 *** | -9.0 *** |
| Malawi | 21.9 *** | 22.6 *** | 26.5 *** | 26.2 *** |
| Mozambique | -6.3 *** | -5.6 ** | -0.2 | 1.9 |
| Niger | -17.5 *** | -16.4 *** | -12.3 *** | -9.9 *** |
| Nigeria | -25.9 *** | -24.4 *** | -21.4 *** | -18.7 *** |
| Sierra Leone | -14.8 *** | -14.2 *** | -12.6 *** | -11.3 *** |
| Tanzania | -10.7 *** | -10.6 *** | -5.9 *** | -5.3 *** |
| Togo | -19.9 *** | -19.6 *** | -18.0 *** | -16.6 *** |
| Uganda | -4.8 *** | -4.4 *** | -4.2 *** | -2.3 * |
| **Facility type (REF = Hospital)** |  |  |  |  |
| Health center | 0.1 | 0.5 | 0.7 | 1.2 |
| Health post | -1.6 | -0.7 | 1.5 | 2.3 * |
| **Managing authority (REF = Private/ NGO)** |  |  |  |  |
| Public | 12.8 *** | 13.2 *** | 11.7 *** | 11.5 *** |
| **Urban/Rural (REF = Urban)** |  |  |  |  |
| Rural | -1.3 * | -0.9 | -0.5 | -0.5 |
| Equipment availability |  | 0.01 ** | 0.01 | 0.01 |
| Infrastructure availability |  | 0.02 *** | 0.02 ** | 0.01 * |
| Absenteeism |  |  | 20.1 *** | 20.2 *** |
| Provider competency |  |  |  | 0.1 *** |
| (Intercept) | 17.8 *** | 14.0 *** | 6.1 *** | 2.3 |
| **R-squared** | 0.2884 | 0.2901 | 0.3359 | 0.3397 |

Significance codes: '\*\*\*' 0.001, '\*\*' 0.01, '\*' 0.05
Note: N = 7236; 1680 facilities excluded due to missing data on caseload. Reasons for missing caseload data include:
- 1) The facility was missing data on the number of staff - 2) The facility was missing data on the number of outpatient visits - 3) The caseload value was set to missing/omitted because it was greater than 200 which was deemed to be unrealistically high

**Supplementary Figure S1a:**
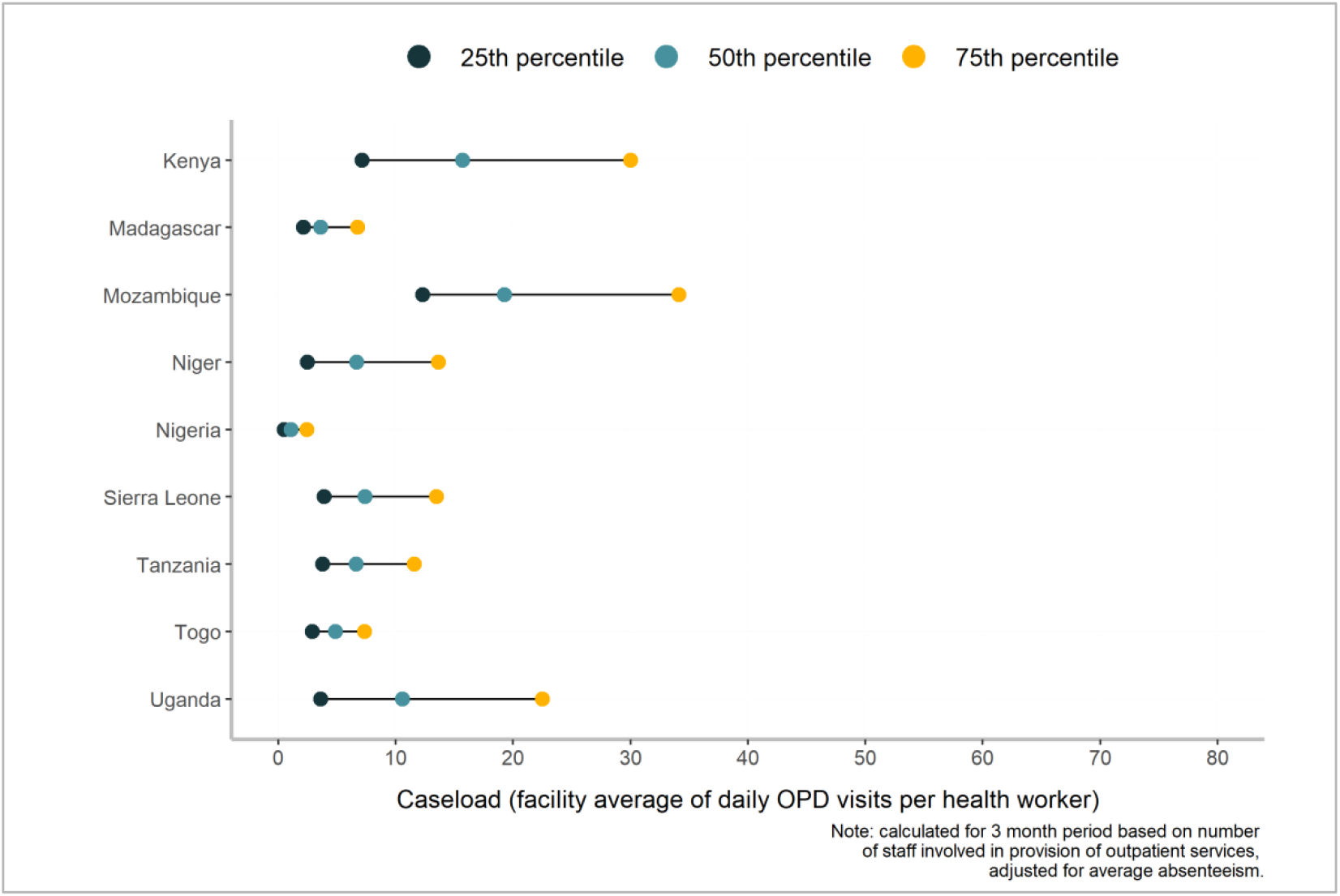
Caseload distribution for health centers and health posts, by country.

**Supplementary Figure S1b:**
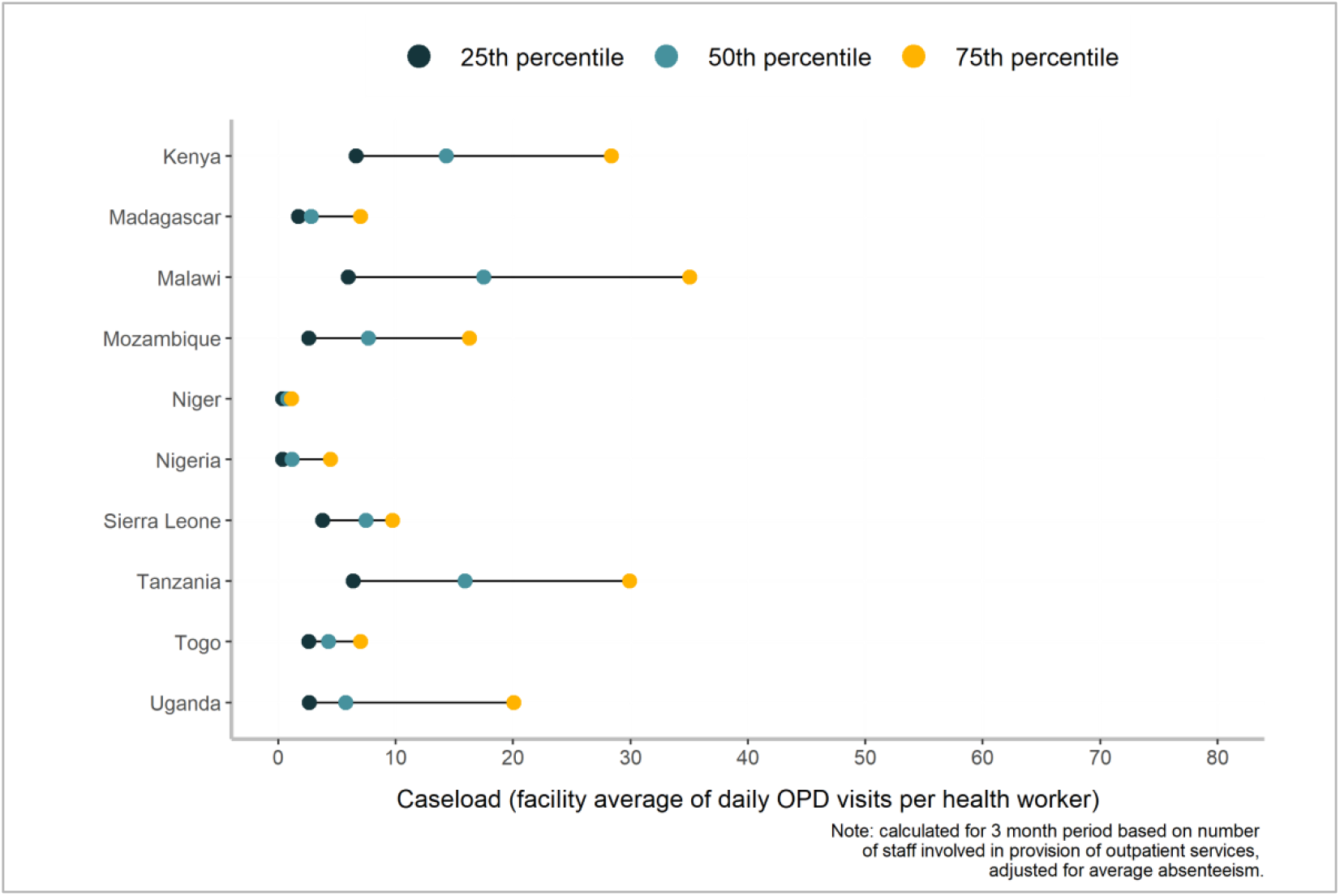
Caseload distribution for hospitals, by country.

**Supplementary Table S13:** Health worker diagnostic accuracy, by country (% and 95% CI)

|  | Kenya | Madagascar | Malawi | Mozambique | Niger | Nigeria | Sierra Leone | Tanzania | Togo | Uganda | All |
| --- | --- | --- | --- | --- | --- | --- | --- | --- | --- | --- | --- |
| National | 66.8<br>(65.6 – 68.0) | 48.1<br>(43.3 - 52.9) | 71.5<br>(69.6 - 73.5) | 58.4<br>(56.0 - 60.7) | 39.9<br>(37.4 - 42.4) | 40.9<br>(38.6 - 43.3) | 49.1<br>(47.0 - 51.2) | 69.2<br>(64.4 – 74.0) | 50.3<br>(45.1 - 55.6) | 56.3<br>(52.7 – 60.0) | 55.1<br>(47.0 - 63.2) |
| Facility type |  |  |  |  |  |  |  |  |  |  |  |
| Hospital | 70.6<br>(66.8 - 74.4) | 54.5<br>(50.3 - 58.7) | 74.3<br>(71.9 - 76.8) | 62.4<br>(59.5 - 65.4) | 44.7<br>(39.6 - 49.9) | 52.0<br>(47.4 - 56.7) | 60.5<br>(54.7 - 66.3) | 71.5<br>(63.3 - 79.6) | 50.1<br>(41.5 - 58.7) | 71.3<br>(60.2 - 82.3) | 61.2<br>(53.7 - 68.7) |
| Health center | 67.1<br>(65.6 - 68.6) | 49.9<br>(43.5 - 56.4) | 67.5<br>(64.6 - 70.5) | 49.6<br>(46.2 - 53.0) | 44.3<br>(40.9 - 47.6) | 31.7<br>(30.4 – 33.0) | 54.1<br>(50.3 – 58.0) | 75.1<br>(65.3 - 84.8) | 56.3<br>(47.9 - 64.7) | 64.2<br>(60.1 - 68.3) | 56.0<br>(46.7 - 65.2) |
| Health post | 64.5<br>(63.5 - 65.4) | 38.5<br>(31.4 - 45.5) | 54.7<br>(47.8 - 61.7) | NA | 30.3<br>(26.4 - 34.3) | 29.3<br>(27.1 - 31.5) | 45.4<br>(42.9 - 47.9) | 56.8<br>(51.7 – 62.0) | 45.3<br>(38.9 - 51.8) | 40.8<br>(37.2 - 44.5) | 45.1<br>(35.9 - 54.2) |
| Managing authority |  |  |  |  |  |  |  |  |  |  |  |
| Private/NGO | 65.4<br>(64.1 - 66.6) | 51.7<br>(45.8 - 57.6) | 67.6<br>(65.4 - 69.9) | 59.7<br>(44.7 - 74.8) | 46.2<br>(40.9 - 51.6) | 52.6<br>(46.2 – 59.0) | 56.8<br>(51.7 – 62.0) | 74.6<br>(64.4 - 84.8) | 54.5<br>(44.2 - 64.8) | 56.2<br>(51.3 - 61.1) | 58.5<br>(52.5 - 64.6) |
| Public | 67.6<br>(65.9 - 69.3) | 46.9<br>(40.9 - 52.9) | 73.0<br>(70.6 - 75.5) | 58.3<br>(55.9 - 60.7) | 39.5<br>(36.9 - 42.1) | 38.4<br>(36.3 - 40.4) | 48.1<br>(45.9 - 50.3) | 66.8<br>(61.4 - 72.1) | 46.9<br>(41.2 - 52.5) | 56.4<br>(51.3 - 61.5) | 54.2<br>(45.5 - 62.9) |
| Urban/Rural |  |  |  |  |  |  |  |  |  |  |  |
| Urban | 67.1<br>(64.5 - 69.7) | 52.1<br>(47.7 - 56.5) | 74.6<br>(71.9 - 77.4) | 67.4<br>(63.4 - 71.3) | 46.5<br>(43.4 - 49.7) | 44.4<br>(40.8 – 48.0) | 54.3<br>(51.2 - 57.4) | 74.2<br>(68.5 - 79.9) | 53.2<br>(45.5 - 60.9) | 64.6<br>(59.4 - 69.9) | 59.8<br>(51.9 - 67.8) |
| Rural | 66.6<br>(65.7 - 67.5) | 45.9<br>(39.0 - 52.8) | 67.4<br>(65.0 - 69.8) | 54.4<br>(51.9 - 56.9) | 34.1<br>(30.7 - 37.4) | 36.5<br>(33.7 - 39.2) | 45.1<br>(42.4 - 47.7) | 56.0<br>(49.6 - 62.5) | 45.7<br>(38.2 - 53.3) | 49.9<br>(46.6 - 53.2) | 50.2<br>(42.1 - 58.2) |
| Health worker cadre |  |  |  |  |  |  |  |  |  |  |  |
| Doctor/<br>Clinical officer | 70.6<br>(68.5 - 72.7) | 57.4<br>(54.1 - 60.6) | 74.6<br>(71.6 - 77.6) | 63.3<br>(60.2 - 66.3) | 57.2<br>(51.2 - 63.1) | 57.6<br>(50.4 - 64.8) | 59.3<br>(49.4 - 69.1) | 72.6<br>(67.0 - 78.2) | 64.3<br>(54.5 - 74.1) | 71.7<br>(67.4 - 75.9) | 64.8<br>(59.9 - 69.8) |
| Nurse /<br>Midwife | 63.0<br>(62.0 - 64.1) | 41.1<br>(33.5 - 48.6) | 70.0<br>(67.4 - 72.5) | 51.7<br>(48.3 - 55.1) | 43.2<br>(39.9 - 46.4) | 44.4<br>(41.1 - 47.8) | 52.0<br>(48.6 - 55.4) | 45.7<br>(39.4 - 52.1) | 46.3<br>(40.8 - 51.8) | 50.2<br>(46.8 - 53.5) | 50.8<br>(44.2 - 57.3) |
| Other worker | 48.5<br>(41.1 - 55.9) | 30.2<br>(16.4 – 44.0) | 54.4<br>(42.2 - 66.6) | 53.7<br>(46.8 - 60.6) | 32.7<br>(29.0 - 36.4) | 30.2<br>(28.8 - 31.7) | 47.5<br>(44.9 - 50.1) | 46.5<br>(37.7 - 55.2) | 39.2<br>(24.1 - 54.4) | 28.7<br>(24.3 – 33.0) | 41.2<br>(33.9 - 48.4) |

**Supplementary Table S14:**
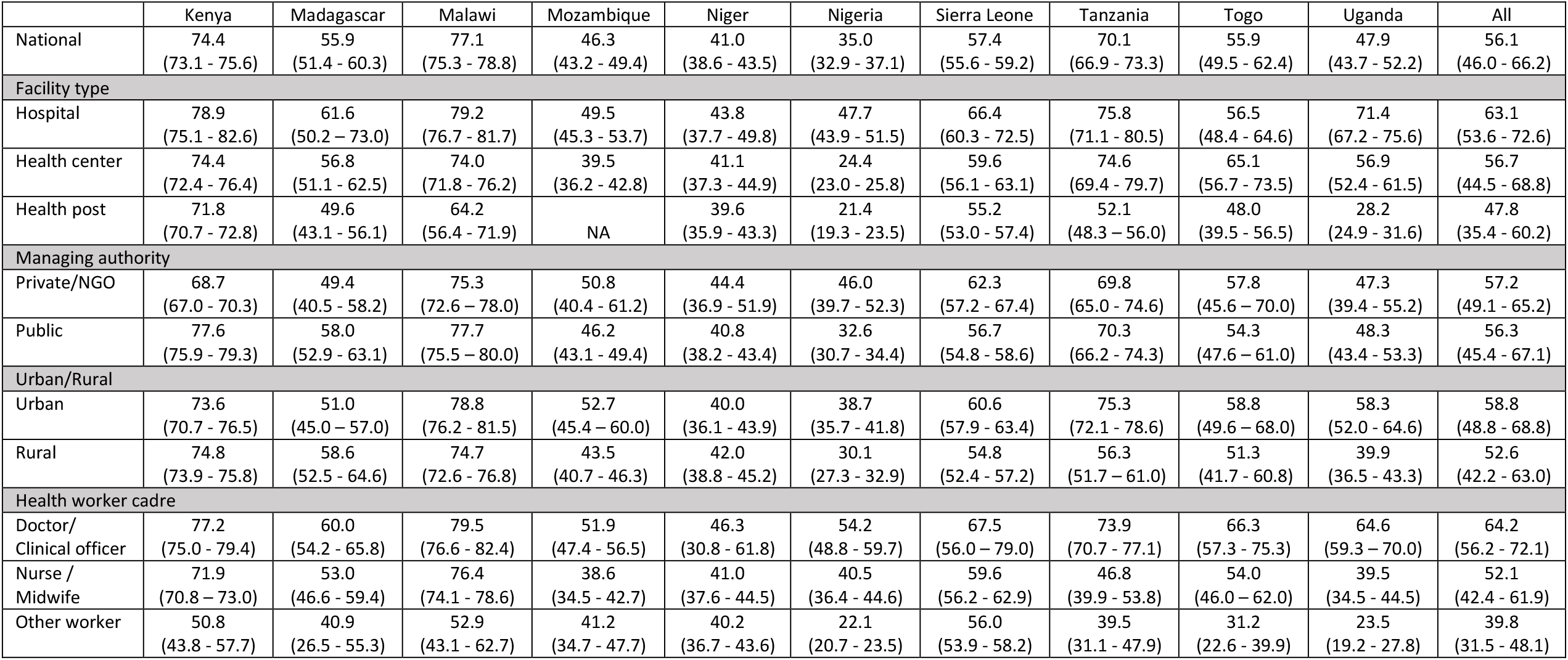
Health worker treatment accuracy, by country (% and 95% CI)

## Footnotes

1 According to the World Development Indicators, Sub-Saharan Africa had 0.2 physicians and just 1 nurse or mid-wife per 1,000 people (2017-2018, the most recent date for which data are available). For South Asia—the region with the next lowest concentration of medical workers—those numbers are 0.8 and 1.5, respectively.

2 For the purposes of this analysis, the term “health worker” refers to all cadres of staff in a facility (e.g., medical staff, nursing staff, community health workers based at the facility). It does not include non-medical staff or community health workers based elsewhere but that report to a health facility. In addition, the term “clinical staff” refers to cadres of health workers who are expected to provide clinical care, more specifically doctors, clinical officers, nurses, and midwives. It does not include cadres of health workers not expected to provide clinical care such as pharmacists and laboratory technicians. For each country, the specific cadres of health workers included as clinical staff differ.

3 Not all clinical staff were assessed for absenteeism (up to a total of 10 health workers were assessed for absenteeism per facility). We explored two alternative methods of adjusting for absenteeism. In the first, we took the total number of clinical staff and reduced that by the number of clinical staff absent. Since not all clinical staff were assessed, this method would undercount absence. In the second, we did the same as in the first, but we restricted the analysis to the subset of facilities where all clinical staff were assessed for absenteeism (i.e., mostly smaller facilities). Results for the sensitivity analyses are presented in ***Supplementary Table S9***. The preferred method (in the main text) allows for a reduction in proportion to the actual absence of clinical staff.

4 Malawi has community level village clinics which report data on service utilization to the health center level. The health center aggregates that data into their monthly reporting forms. This may have led to an inflation in the number of outpatient services delivered at the primary level. While there is evidence of low health worker density and high service utilization as an ongoing problem in Malawi, we could not confirm the validity of the caseload data and therefore have excluded it from the analysis.

5 Hospital includes hospitals of all levels (district, regional, national)

6 Health center includes facilities designed to run as outpatient clinics and may include inpatient treatment or a maternity ward. These are facilities characterized by higher level-staff with the ability to diagnose and treat a range of health conditions and responsibility for a larger catchment area (e.g., health center, level 2 basic health center, integrated health center).

7 Health post includes facilities designed to run as outpatient clinics treating common diseases and offering antenatal care (e.g., dispensary, level 1 basic health center, health hut).

8 The combined nurse/midwife category was created considering the different staff categorization between countries and to allow for cross country comparability. The “other health workers” category was created for health workers not expected to provide clinical care such as laboratory technicians, pharmacists, public health officers, and health records information officers.

9 Survey weights were calculated for facilities as the inverse of the probability of selection for the facility inventory. Survey weights were calculated for health workers separately for absenteeism and health worker competency. Absenteeism weights were calculated as the inverse of the probability of selection for the absenteeism assessment based on the total number of medical staff recorded on the staff roster times the facility weight. Competency weights were calculated as the inverse of the probability of selection for the competency vignette based on the total number of medical staff who see outpatients recorded on the staff roster times the facility weight. Facility level sample weights were unavailable for Mozambique so unweighted facility level results are reported.

10 For the caseload regression, mean competency scores were missing for 11 facilities (0.2% of data). These missing values were imputed using the country average mean competency score.

11 We have chosen not to disaggregate results for facilities with 0 versus 1 clinical health worker as both situations highlight a staffing challenge of concern.

12 Zero clinical staff does not mean zero staff. Some facilities have no staff who are classified as those authorized to treat patients; but in practice, some other staff may treat patients.

13 For more on provider competency, see section 3.4.

